# Genome-Wide Association and Population-Tailored Polygenic Risk for Parkinson’s Disease in Taiwan

**DOI:** 10.64898/2026.01.14.26344088

**Authors:** Yung-Tsai Chu, Yu-An Su, Chin-Hsien Lin, Chun-Hwei Tai, Yih-Ru Wu, Chien-Tai Hong, Yu-Wei Chen, Mong-Hsun Tsai, John Hardy, Kin-Ying Mok, Ruey-Meei Wu, East Asian Parkinson Disease Genomics Consortium, Global Parkinson’s Genetics Program

## Abstract

Parkinson’s disease genetics remain under-characterized in East Asians. We recruited a Taiwanese case–control cohort (2,245 PD; 2,147 controls), genotyped on the Illumina NeuroBooster Array, and imputed 7.6 million variants using the Taiwan Biobank reference. Logistic-regression GWAS identified genome-wide significant signals at *SNCA* and *MCCC1*; we also observed suggestive associations at *GCH1, PPARGC1A* and *GALNT13*. Haplotype analyses delineated an East Asian *SNCA* risk haplotype and confirmed effects at *LRRK2* p.G2385R and p.Rorg8P. Moreover, we have discovered, for the first time, a gene dosage effect of *LRRK2* Asian variants. A European-derived PRS (Nalls 2019) showed modest discrimination in our cohort (AUC 0.590); incorporating Asian and Taiwan-specific variants increased the AUC to 0.615 (DeLong p<0.001). Our data define the genetic architecture of PD in Taiwan, highlight both shared and population-specific risk, and demonstrate that tailoring polygenic risk scores to ancestry-specific genetic structure improves risk stratification.

## Introduction

Parkinson’s disease (PD) is the second most common neurodegenerative disorder worldwide.^1^ It is characterized clinically by tremor, rigidity, bradykinesia, and postural instability, and neuropathologically by selective dopaminergic neuron loss in the substantia nigra with Lewy bodies composed primarily of α-synuclein.

Although a minority of PD cases arise from rare, high-penetrance variants, most cases are polygenic and multifactorial. Common variants of modest effect collectively account for a meaningful portion of PD heritability; the largest European-ancestry meta-GWAS to date identified 90 independent risk signals explaining an estimated 16–36% of the heritable risk.^2^ Yet the field’s understanding of PD genetics remains disproportionately derived from European cohorts, with fewer large-scale studies in East Asian and Taiwanese populations.^3^

Both clinical features and genetic risk architectures show ancestry-related differences.^4^ Asian patients, for example, have lower reported rates of motor complications such as dyskinesia than European patients, and several PD risk alleles differ in frequency and effect size across populations. Notably, the *LRRK2* p.G2019S founder variant is enriched in European and certain West Asian groups, whereas *LRRK2* p.G2385R and p.R1628P are established risk alleles in individuals of East Asian ancestry.^5–7^ At *SNCA*, ancestry-specific patterns have also been described: association signals and local haplotype structure vary by population^8,9^, and distinct rs356182-centered haplotypes show different relationships to risk in Latino and European cohorts, highlighting locus complexity relevant to non-European populations as well. In GBA1, recent GWAS in African cohorts have likewise identified novel PD risk variants among individuals of African ancestry.^10^

These differences have direct implications for risk prediction^11^. Cross-ancestry evaluations consistently show that polygenic risk scores (PRS) trained on European data underperform in non-European populations, arguing for population-tailored models. In response to this imbalance, the East Asian Parkinson Disease Genomics Consortium (EAPDGC) was formed to coordinate multi-center efforts across the region.^12^ We undertook the first Taiwan-wide PD GWAS to (i) map PD risk loci and related haplotypes in a Taiwanese cohort, (ii) benchmark effect sizes against European and pan-East Asian studies, and (iii) construct a Taiwan-optimized PRS to assess improvements in predictive performance over European- and Asian-derived baselines.

## Results

### Cohort demographics

After quality control, the final cohort comprised **2,251** individuals with Parkinson’s disease (PD) and **2,268** controls (Supplementary Table S1 lists recruitment by site). Cases were slightly older at examination than controls (65.4 ± 11.2 vs. 63.5 ± 8.7 years) and more often male (51.5% vs. 44.4%). The mean age at PD onset was **60.5 ± 11.7** years. Key characteristics are summarized in Table 1. Age and sex were included as covariates in all association models. Population structure was evaluated by principal component analysis (PCA) using the 1000 Genomes Phase 3 reference panel; study participants clustered with the East Asian (EAS) superpopulation (CHB/CHS/JPT/KHV/CDX) (Supplementary Fig. S1).

**Table 1.**
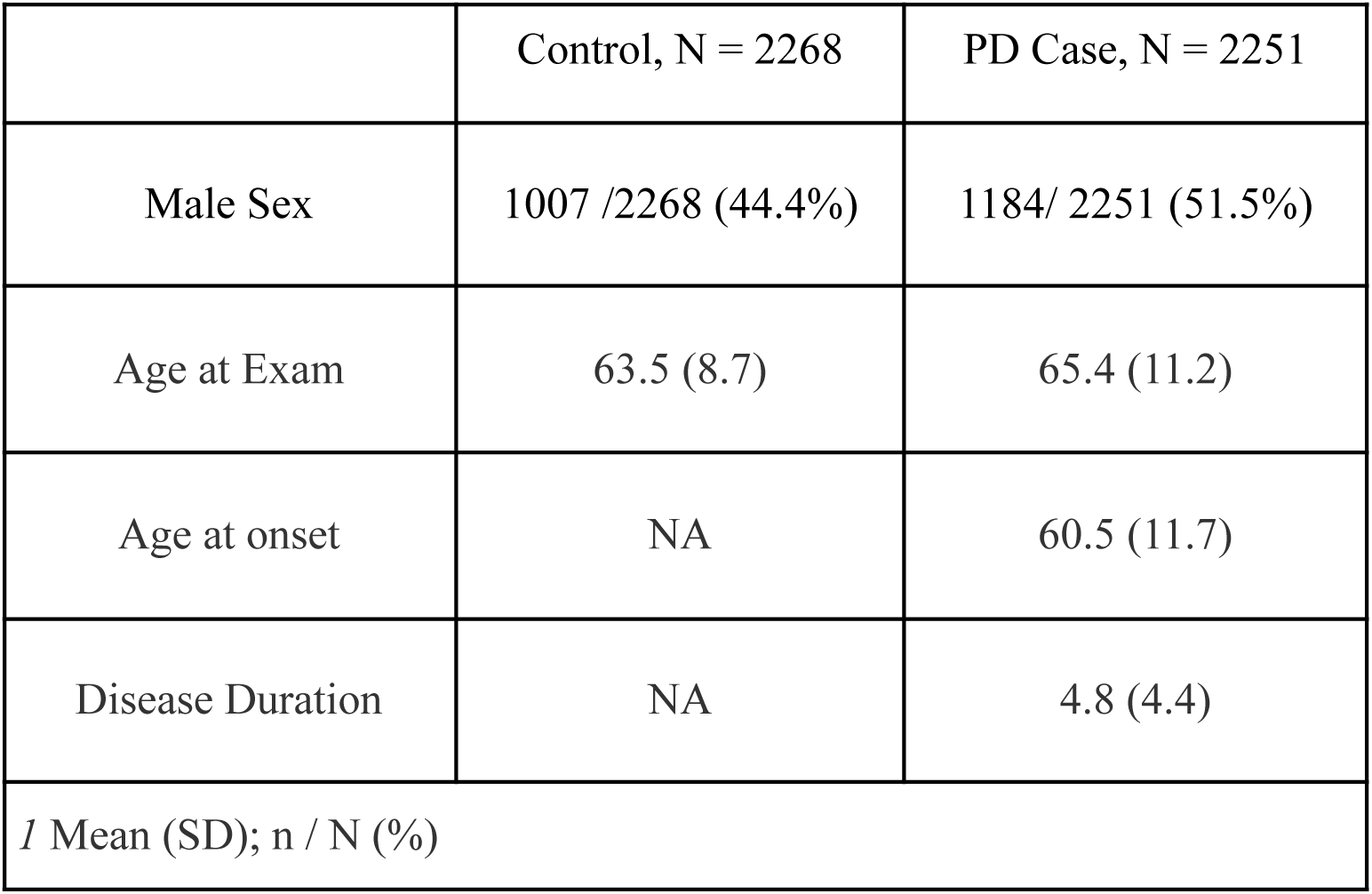
Demographics of the study cohort after quality control. Summary of age, sex, and case–control distribution of participants included in the genome-wide association study after quality control. The detailed number of participants contributed by each recruitment site is provided in **Supplementary Table 1**.

|  | Control, N = 2268 | PD Case, N = 2251 |
| --- | --- | --- |
| Male Sex | 1007 /2268 (44.4%) | 1184/ 2251 (51.5%) |
| Age at Exam | 63.5 (8.7) | 65.4 (11.2) |
| Age at onset | NA | 60.5 (11.7) |
| Disease Duration | NA | 4.8 (4.4) |
| *I* Mean (SD); n / N (%) |  |  |

### Genome-wide association analysis (Fig. 1; Tables 2–3)

For the GWAS, after imputation and model-covariate completeness filters, **2,245** PD cases and **2,147** controls of Taiwanese ancestry remained, contributing ∼**7.6 million** high-quality variants. The quantile–quantile plot showed minimal test-statistic inflation (λ_GC_=**1.02**), indicating adequate control of population structure (Supplementary Fig. S2).

**Figure 1.**
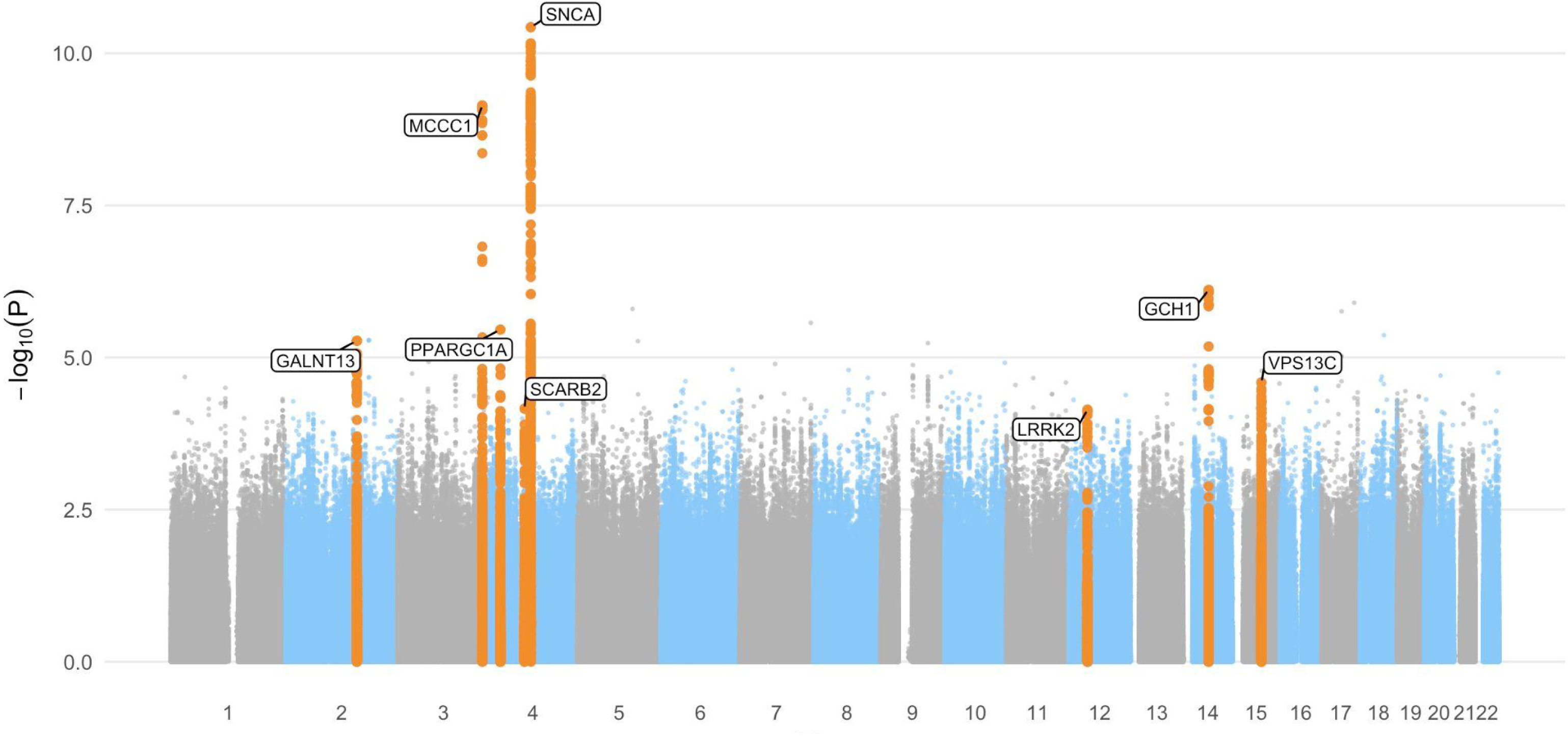
Genome-wide association results for Parkinson’s disease in the Taiwanese cohort. The Manhattan plot displays –log_10_p values for approximately 7.6 million variants after quality control and imputation. The red dashed line denotes the threshold for genome-wide significance (p = 5 × 10^-8^), and the blue dashed line indicates the threshold for suggestive significance (p = 1 × 10^-5^). Established PD risk loci, including *SNCA*, *MCCC1*, and *GCH1*, were successfully replicated. Additional signals in *PPARGC1A* and *GALNT13* emerged as potential novel associations specific to the Taiwanese population. Although not exceeding the suggestive threshold, loci in *VPS13C*, *LRRK2* (p.G2385R), and *SCARB2* also demonstrated supportive evidence consistent with prior reports, suggesting potential relevance in East Asian populations.

**Table 2:**
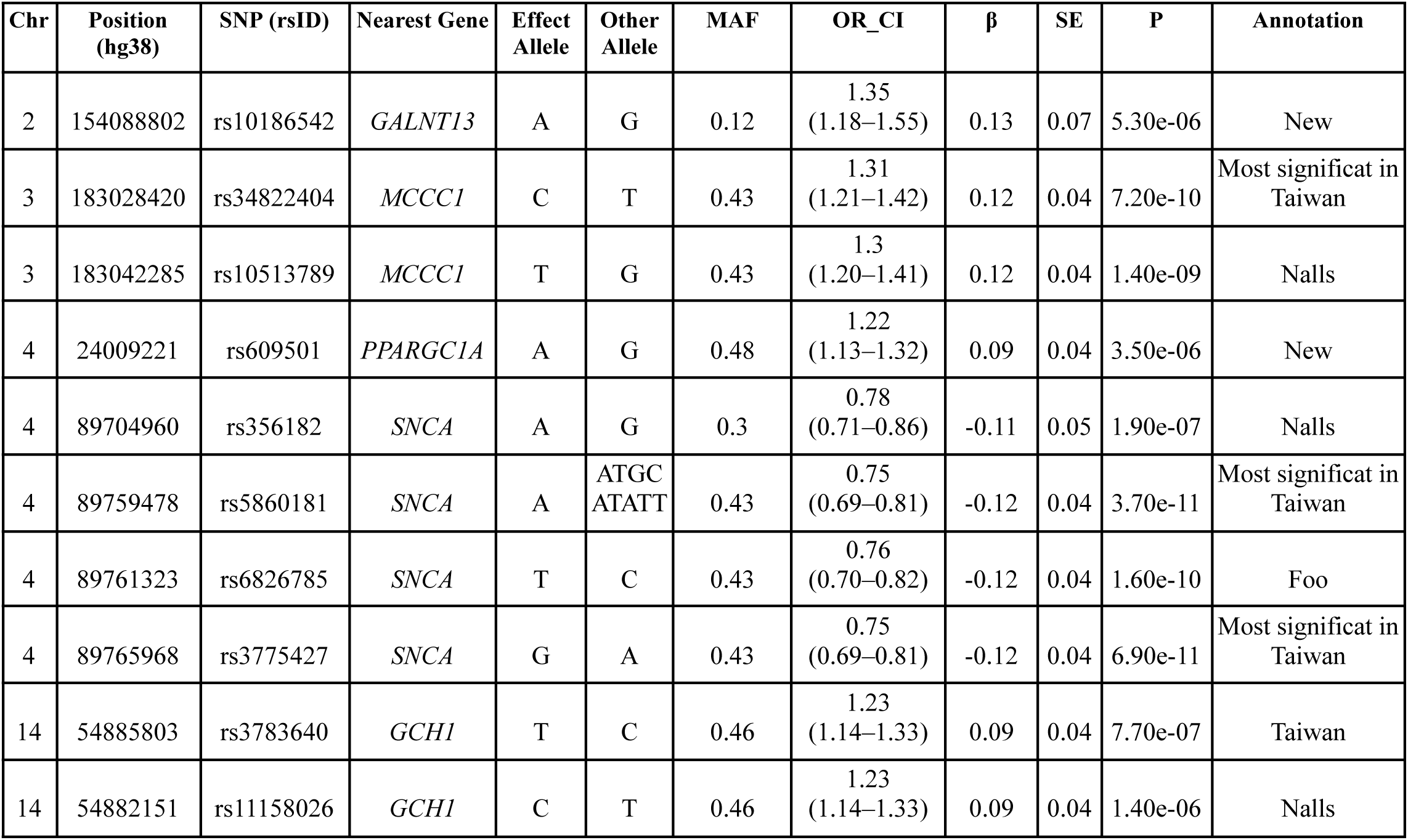
Top Genetic Risk Loci Associated with PD in Taiwan.

**Table 3:**
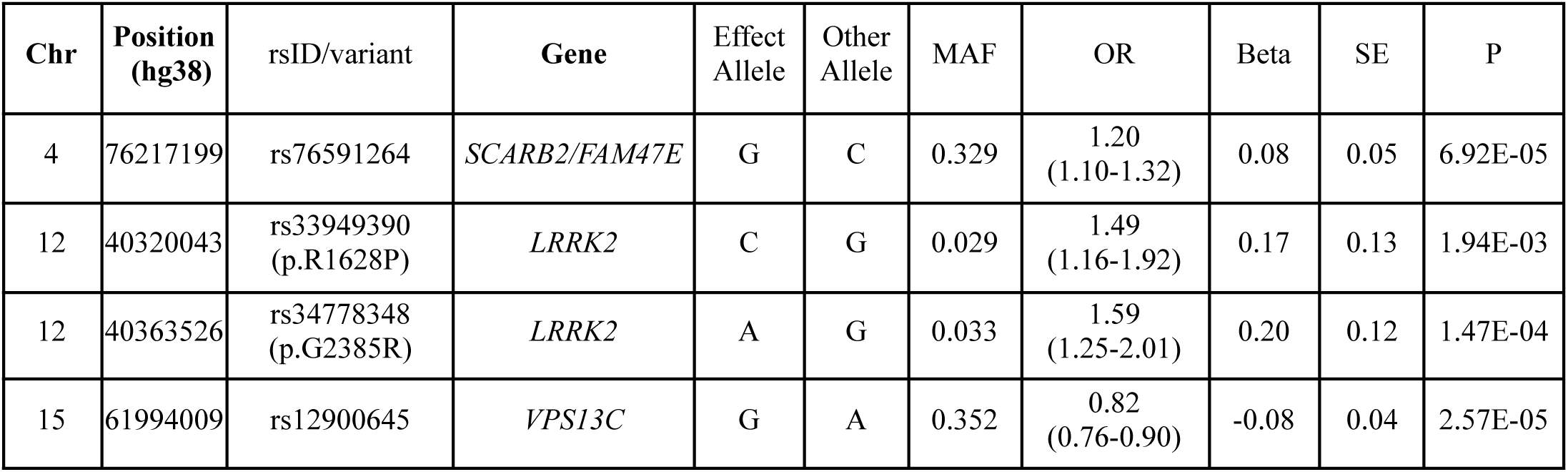
Replication of established Parkinson’s disease loci and targeted look-up in the Taiwanese cohort.

| Chr | Position (hg38) | rsID/variant | Gene | Effect Allele | Other Allele | MAF | OR | Beta | SE | P |
| --- | --- | --- | --- | --- | --- | --- | --- | --- | --- | --- |
| 4 | 76217199 | rs76591264 | *SCARB2/FAM47E* | G | C | 0.329 | 1.20<br>(1.10-1.32) | 0.08 | 0.05 | 6.92E-05 |
| 12 | 40320043 | rs33949390<br>(p.R1628P) | *LRRK2* | C | G | 0.029 | 1.49<br>(1.16-1.92) | 0.17 | 0.13 | 1.94E-03 |
| 12 | 40363526 | rs34778348<br>(p.G2385R) | *LRRK2* | A | G | 0.033 | 1.59<br>(1.25-2.01) | 0.20 | 0.12 | 1.47E-04 |
| 15 | 61994009 | rs12900645 | *VPS13C* | G | A | 0.352 | 0.82<br>(0.76-0.90) | -0.08 | 0.04 | 2.57E-05 |

The Manhattan plot (Fig. 1) highlighted signals at ***SNCA*** and ***MCCC1***, with additional support at ***GCH1*** and several suggestive loci (P<1×10^-5^). The most significant associations localized to established PD risk regions—*SNCA*, *MCCC1*, and *GCH1*. Beyond these, we observed suggestive associations at *PPARGC1A* and *GALNT13*, nominating potentially Taiwan-specific risk variants. Top results are summarized in Table 2 and Supplementary Table S2. Detailed regional association plots are provided in Supplementary Fig. S3.

The lead association was **rs5860181** within intron 4 of *SNCA* (odds ratio [OR]=**0.75**, 95% CI 0.68–0.84, P=**3.7×10**^-11^)**(Fig. 2)**. A prominent signal was detected at **MCCC1** (rs34822404; OR=**1.31**, 95% CI 1.19–1.44, P=**7.2×10**^-10^), consistent with prior East Asian reports. At ***GCH1***, we replicated association at **rs3783640** (OR=**1.23**, 95% CI 1.09–1.39, P=**7.7×10^-7^**), which is in linkage disequilibrium with the previously reported rs11158026 signal. (Supplementary Fig. S4)

**Figure 2.**
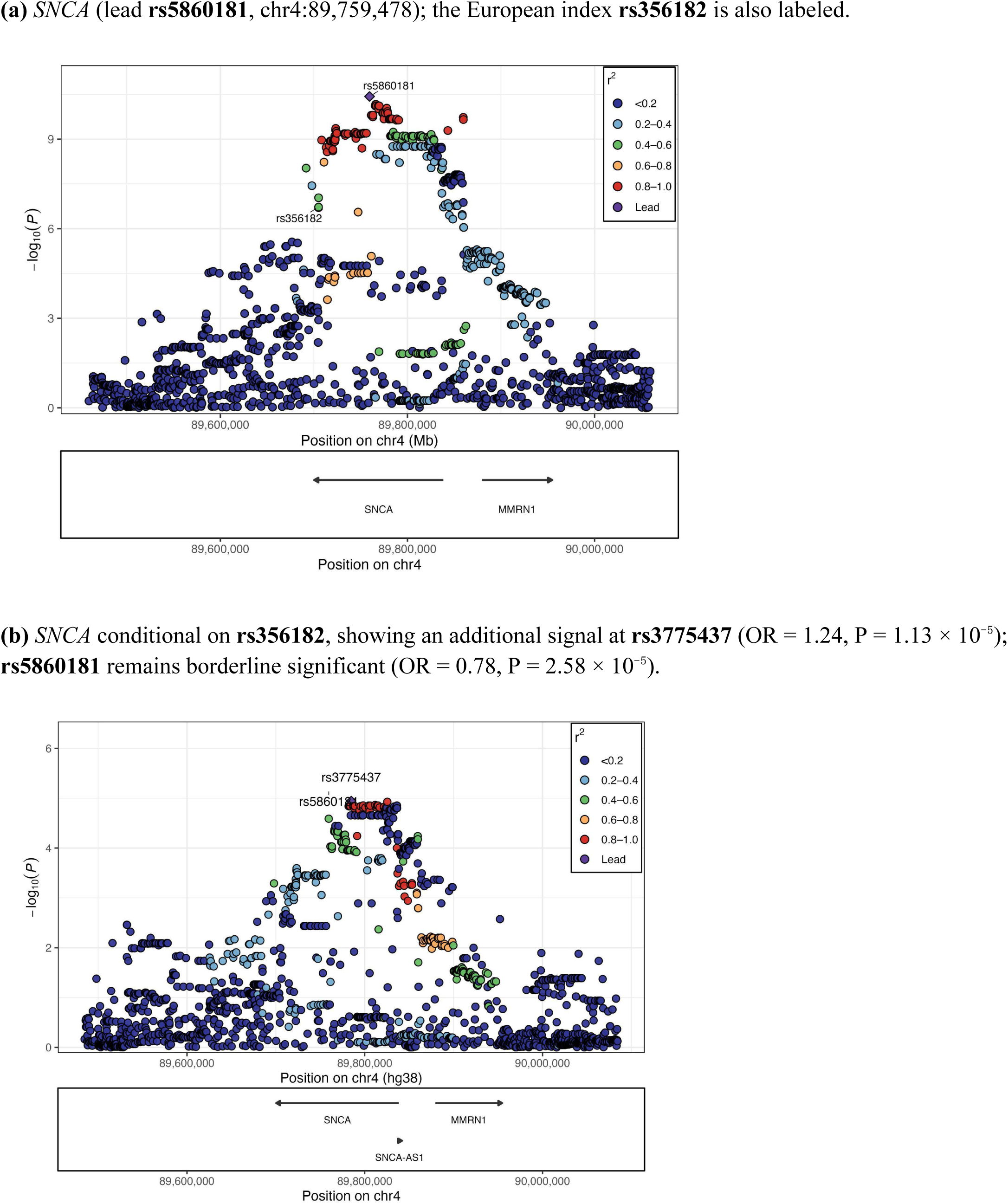
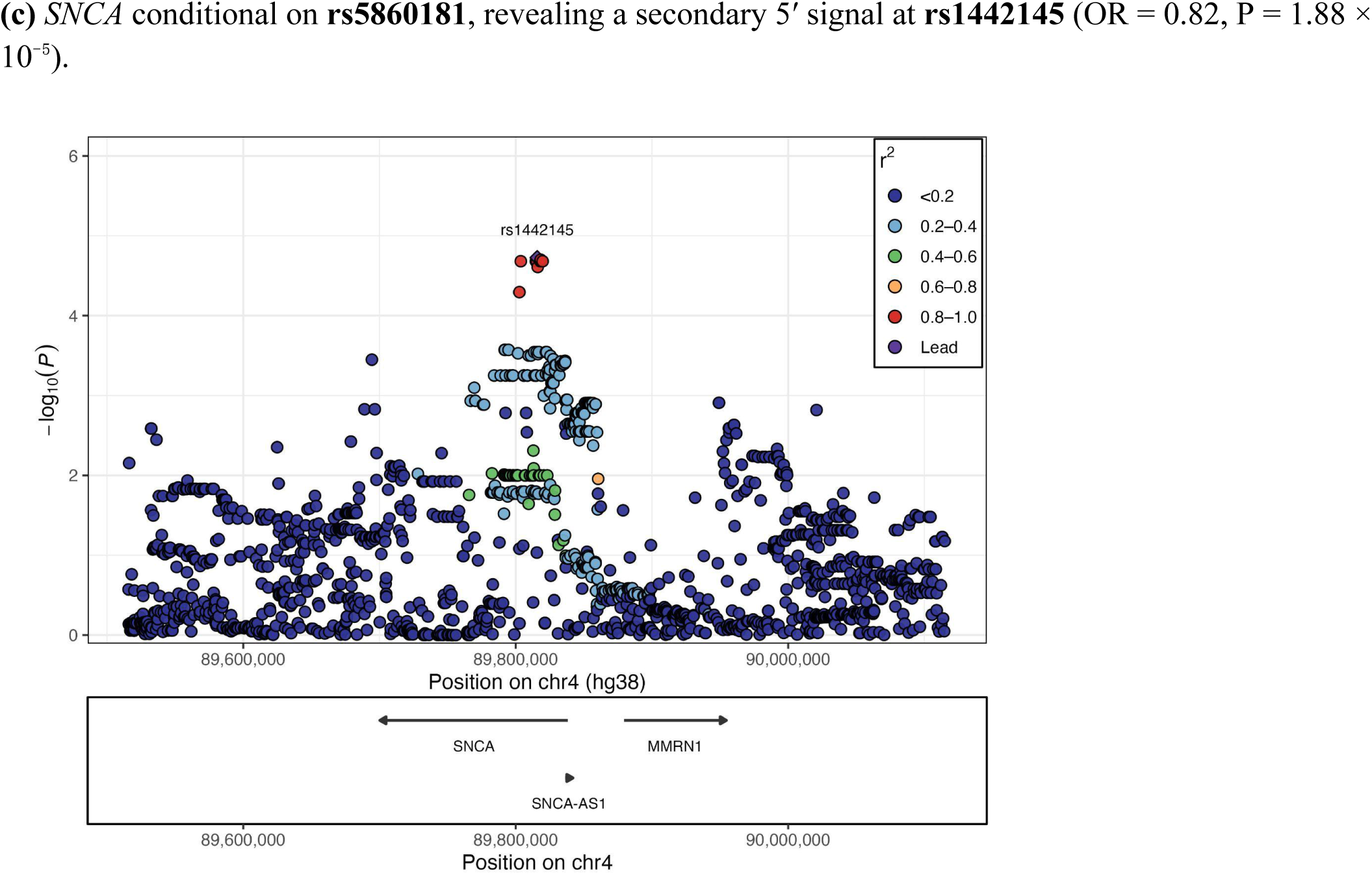
*SNCA* regional association. LocusZoom-style plots display –log₁₀(P) by genomic position across ±300 kb of the indicated lead SNP. Linkage disequilibrium (LD; r²) is computed from cohort controls; the lead SNP is shown in purple and other variants are colored by r² with the lead.

Among suggestive loci, ***PPARGC1A*** (rs609501; OR = 1.22, 95% CI 1.08–1.38, P = 3.5 × 10^-6^) and ***GALNT13*** (rs10186542; OR = 1.35, 95% CI 1.01–1.45, P = 5.3 × 10^-6^) have not, to our knowledge, been reported in prior PD GWAS and warrant follow-up. We also observed the expected positive effects for ***LRRK2*** p.G2385R and p.R1628P (Table 3). Finally, variants within ***VPS13C*** and ***SCARB2*** that were not in high LD with previously reported index variants from Nalls et al. or Foo et al. (Supplementary Fig. S3 e-f) suggest potentially Taiwan-specific signals. Of note, the protective VPS13C signal at rs12900645 (chr15:61,994,009; hg38) lies within the putatively protective haplotype p.R153H–p.I398I–p.I1132V–p.Q2376Q (chr15:61,920,582–62,023,836) reported by Rudakou et al.^13^ In Taiwanese controls, linkage disequilibrium between **rs12900645** and **p.I1132V** was modest (**r² ≈ 0.44**), indicating only partial tagging of the Rudakou haplotype. Across the four coding/synonymous variants reported by Rudakou et al., point estimates were protective but not statistically significant (e.g., **p.I1132V OR = 0.91, P = 0.08**). These findings are consistent with a protective background spanning **chr15:61,920,582–62,023,836**; however, in the Taiwanese cohort the **intronic rs12900645** is the **lead marker** of the protective effect in *VPS13C*.

### *SNCA* conditional analysis (Fig. 2)

The regional association at *SNCA* identified **rs5860181** (intron 4) as the lead variant (OR = 0.75, **P = 3.7 × 10**^-11^), whereas the European index **rs356182** (3′ UTR) was less significant in Taiwanese cohort (OR = 0.78, **P = 1.9 × 10^-7^**) (Fig. 2a). Conditioning on **rs356182** revealed a residual intron-4 signal at **rs3775437** (OR = 1.24, **P = 1.13 × 10^-5^**) and **rs5860181** remained borderline significant (OR = 0.78, **P = 2.58 × 10^-5^**), indicating partial independence between the intron-4 (Taiwan-enriched) signal and the rs356182 haplotype (Fig. 2b). Conversely, conditioning on **rs5860181** revealed a secondary 5′ signal at **rs1442145** (OR = 0.82, **P = 1.88 × 10^-5^**), while **rs356182** was no longer significant (OR = 0.94, **P = 0.33**; Fig. 2c).

### Genotype analyses of *LRRK2* risk alleles (Table 4)

The combined carrier rate of East Asian *LRRK2* risk variants was **10.8%** in controls and **16.3%** in PD. Relative to noncarriers, individuals carrying **one** variant (either p.G2385R or p.R1628P) had **OR = 1.49** (95% CI 1.23–1.79; **P = 4.1 × 10^-5^**). Carriers of **two** risk variants (p.G2385R homozygotes, p.R1628P homozygotes, or p.G2385R/p.R1628P compound heterozygotes) had **OR = 4.30** (95% CI 1.59–11.6; **P = 0.004**) versus noncarriers and **OR = 2.89** (95% CI 1.06–7.91; **P = 0.039**) versus single-variant carriers, consistent with a gene-dosage effect.

**Table 4.** Distribution of East Asian *LRRK2* risk genotypes and associated PD odds. Odds ratios from logistic regression adjusted for age, sex, and top 15 PCs; reference = noncarriers. “Double-variant” includes G2385R/G2385R, R1628P/R1628P, and G2385R/R1628P compound heterozygotes.

| Table 4A |  |  |  |  |
| --- | --- | --- | --- | --- |
| LRRK2 risk variant number | 0 | 1* | 2** | *LRRK2* Carrier Rate |
| Control | 2047 | 216 | 5 | 10.8% |
| PD | 1935 | 297 | 19 | 16.3% |
| Onset age | 60.75 ± 11.19 | 59.17 ± 11.25 | 62.00 ± 9.38 | One-way ANOVA: p value 0.07 |
| * either *LRRK2* G2385R or R1628P one variant carrier |  |  |  |  |
| ** double variants : G2385R homozygous, R1628P homozygous or G2385R + R1628P compound heterozygous |  |  |  |  |
| Table 4B |  |  |  |  |
|  | Odds Ratio | 95% CI | p value |  |
| single variant vs. non-carrier | 1.49 | 1.23-1.79 | 4.1E-05 |  |
| double variants vs. non-carrier | 4.29 | 1.59-11.6 | 0.004 |  |
| double variants vs. single variant | 2.89 | 1.06-7.9 | 0.039 |  |

Age at onset (mean ± SD) did not differ significantly across groups: noncarriers **60.75 ± 11.19** years, one-variant carriers **59.17 ± 11.25** years, and two-variant carriers **62.00 ± 9.38** years. In an exploratory subgroup comparison, **compound heterozygotes (p.G2385R/p.R1628P)** showed a numerically earlier onset (**55.6 ± 5.8** years) versus noncarriers (**60.75 ± 11.19** years), a trend that did not reach significance (**t-test P = 0.06**); this analysis is limited by small sample size.

### Haplotype structure at *SNCA*, *LRRK2*, and *GCH1* (Fig. 3)

To probe ancestry-specific architecture, we performed regional haplotype analyses for *SNCA*, *LRRK2*, and *GCH1*.

**Figure 3.**
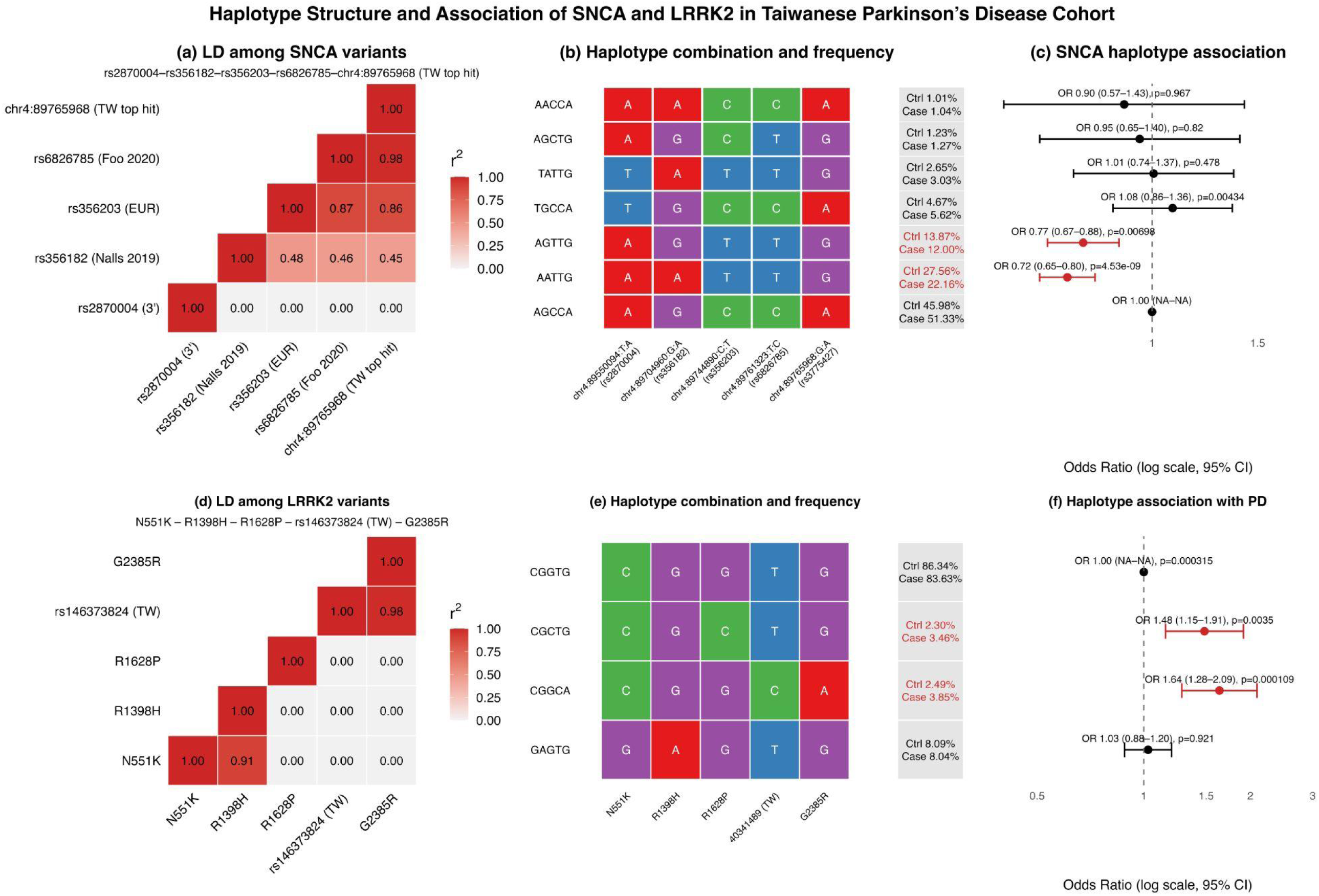
Haplotype structure and association of SNCA and LRRK2 in the Taiwanese Parkinson’s disease cohort. **(a)** Linkage disequilibrium (LD) heatmap (r²) among five representative *SNCA* variants (rs2870004, rs356182, rs356203, rs6826785, rs3775427) computed in controls. **(b)** Case–control frequencies of common *SNCA* haplotypes (allele order as in panel a); bases are color-coded by nucleotide. Protective haplotypes **AGTTG** and **AATTG** are highlighted. **(c)** Forest plot of *SNCA* haplotype associations (odds ratio [OR] with 95% confidence interval [CI]; log scale) with corresponding P values; **AGTTG** and **AATTG** show reduced PD risk. **(d)** LD heatmap (r²) for five *LRRK2* missense/region-specific variants—N551K (rs7308720), R1398H (rs7133914), R1628P (rs33949390), chr12:40341489, and G2385R (rs34778348)—illustrating an East Asian–patterned LD structure. **(e)** *LRRK2* haplotype composition and case–control frequencies (allele order as in panel d). **(f)** Forest plot of adjusted *LRRK2* haplotype associations (OR, 95% CI, P). Haplotypes **CGGCA** and **CGCTG** exhibit protective effects relative to the reference **CGGTG**. **Abbreviations:** LD, linkage disequilibrium; OR, odds ratio; CI, confidence interval. **Analysis details:** LD was estimated with PLINK v1.9 (–r²) in controls. Haplotypes were inferred by EM (PLINK) and tested for association using the R package **haplo.stats**, with models adjusted for age, sex, and the first five principal components.

#### SNCA

The lead variant **rs3775427** (intron 4) resides on an East Asian–enriched haplotype distinct from the European **rs356182**–centered configuration. Using 1000 Genomes East Asian reference data via LDlink, **rs3775427**—the top *SNCA* signal in Taiwan—was tightly correlated with **rs6826785** (D′ = 0.996, r² = 0.988) and only partially correlated with **rs356182** (D′ = 0.83, r² = 0.469). Haplotype analysis of five representative variants (rs2870004, rs356182, rs356203, rs6826785, rs3775427) identified two common configurations, **AGTTG** and **AATTG**, that were associated with reduced PD risk (Fig. 3a–c). Notably, both protective haplotypes carry the **rs3775427-G** allele, whereas the **rs356182** allele differs between them (A vs. G), consistent with **rs3775427-G** (or variants on its tight LD background) tagging the protective effect. These findings support population-specific LD blocks at *SNCA*, with the Taiwanese pattern aligning more closely with other East Asian cohorts than with Europeans.

#### LRRK2

We evaluated five missense/region-specific variants—p.**N551K** (rs7308720), p.**R1398H** (rs7133914),p. **R1628P** (rs33949390), an intronic variant at chr12:40341489, and East Asian–specific p.**G2385R** (rs34778348). LD patterns indicated three largely independent variant groups (N551K–R1398H; R1628P; G2385R). Haplotype reconstruction followed by adjusted logistic regression identified **CGGCA** and **CGCTG** as **risk** haplotypes relative to the reference **CGGTG**, whereas the N551K–R1398H combination did not show a significant protective effect (Fig. 3d–f). Further principal component analysis (Fig. 4) shows that carriers of both *LRRK2* Asian variants cluster mainly within CHS (Southern Han Chinese). p.R1628P carriers are additionally observed in CDX (Chinese Dai) and KHV (Kinh Vietnamese), suggesting a more southern extension, whereas p.G2385R carriers are also seen in CHB (Han Chinese in Beijing) and JPT (Japanese), indicating a more northeastern distribution.

**Figure 4.**
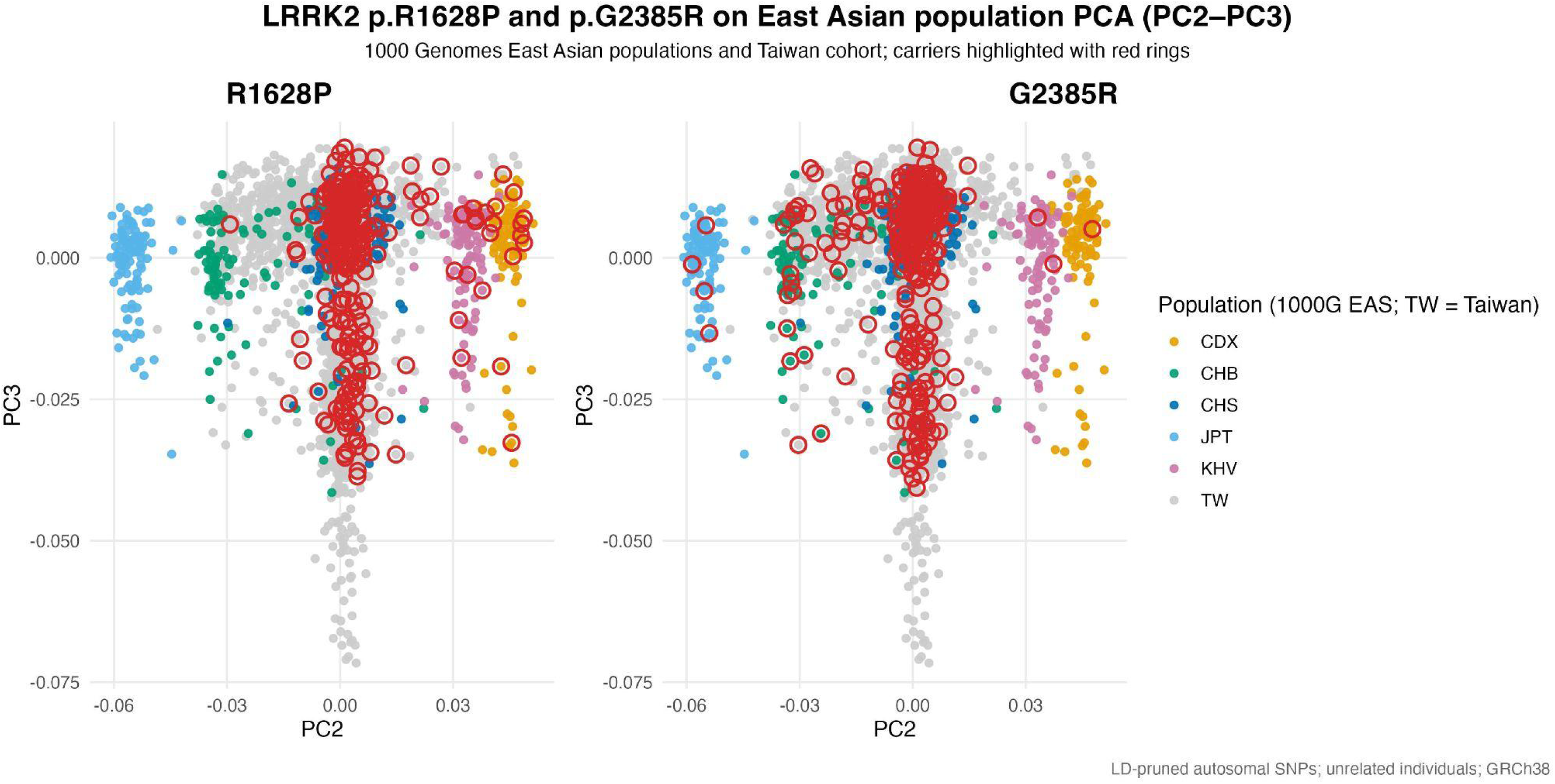
LRRK2 p.R1628P and p.G2385R on East Asian population PCA (PC2–PC3) Principal component analysis using LD-pruned autosomal SNPs (GRCh38) from the 1000 Genomes East Asian populations (CDX, CHB, CHS, JPT, KHV) and a Taiwanese cohort (TW). Points are colored by population; carriers of LRRK2 p.R1628P (left) and p.G2385R (right) are indicated by red hollow circles. Related individuals were removed prior to PCA.

#### GCH1

In Taiwan, **rs3783640** is in LD with **rs11158026** reported previously; both are only in partial LD with **rs841** (r²≈0.31; Supplementary Fig. 2). Haplotype analyses indicated that configurations carrying **rs3783640** and **rs11158026** are associated with PD risk, whereas **rs841** alone was not significantly associated, suggesting that prior protective signals attributed to rs841 may be mediated by rs11158026/rs3783640.

Together, these analyses illustrate that while *SNCA*, *LRRK2*, and *GCH1* are globally implicated in PD, internal LD and haplotypic configurations differ by ancestry, underscoring the value of population-tailored analyses.

### Cross-ancestry comparison (Fig. 5; Supplementary Table S3)

We compared effect sizes in this Taiwanese GWAS with those from a large European meta-analysis (Nalls 2019) and a pan-East Asian GWAS (Foo 2020). Scatter plots of log-odds ratios showed a **modest but significant** correlation with European estimates (Pearson’s r=**0.67**, P=**2.3×10^-^**^11^), with over half of variants not reaching significance in our cohort (green points in Fig. 3a). In contrast, effects were **strongly concordant** with East Asian estimates (r=**0.96**, P=**7.5×10^-6^**), reflecting shared risk architecture within East Asia. Notably, the ***MCCC1*** risk allele exhibited a larger effect in the Taiwanese cohort (OR=**1.32**) compared with European cohorts (OR≈**1.1**), suggesting population-specific modulation of genetic risk.

**Figure 5.**
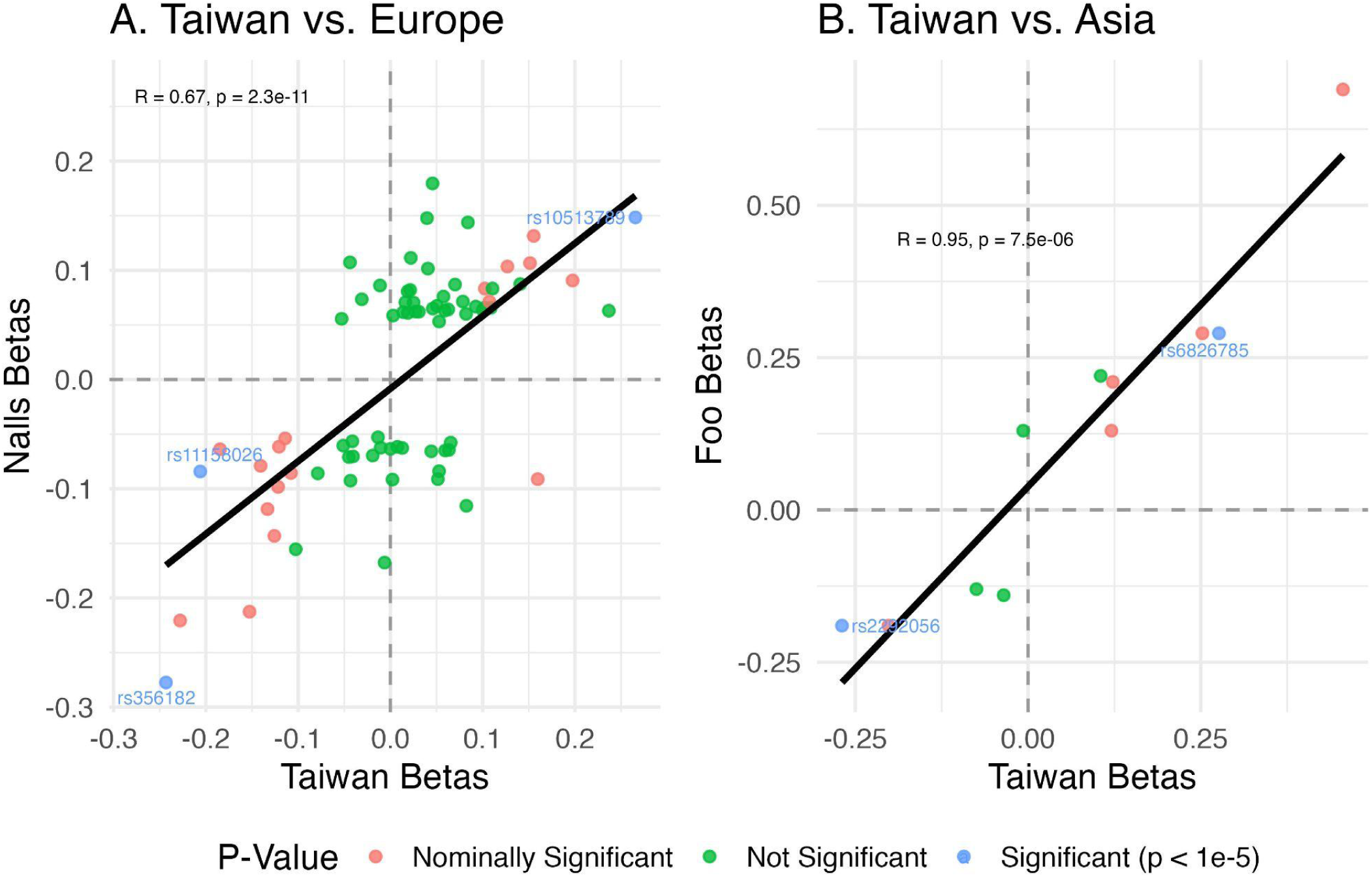
Replication of genome-wide significant variants from Nalls et al. (2019) and Foo et al. (2020). Scatter plots compare the effect sizes (log odds ratios) of PD-associated variants identified in previous GWAS with those observed in the Taiwanese cohort. For detailed statistics, please refer to Supplementary Table 4. Green points represent variants that were not statistically significant in the current dataset. A modest correlation was observed with the European meta-analysis by Nalls et al. (2019) (R = 0.67, p = 2.3 × 10⁻¹¹), reflecting limited cross-ancestry transferability of risk alleles. In contrast, effect sizes showed a strong positive correlation with the pan-Asian GWAS by Foo et al. (2020) (R = 0.96, p = 7.5 × 10⁻⁶), indicating a high degree of genetic concordance and shared PD risk architecture within East Asian populations.

### Polygenic risk scores and population-specific improvement (Fig. 6)

We evaluated three PRS models. Using **76** variants from the European-based GWAS (Nalls 2019), the baseline PRS achieved **AUC 0.590** (95% CI 0.574–0.607). Augmenting with **11** independent Asian variants from Foo 2020 (excluding 9 overlapping loci) improved AUC to **0.606** (95% CI 0.590–0.623). Further inclusion of Taiwan-specific variants (***LRRK2* p.G2385R**, ***LRRK2* p.R1628P**, ***VPS13C* rs12900645**, ***SCARB2/FAM47E* rs76591264**) yielded **AUC 0.615** (95% CI 0.599–0.632). Scoring weights are provided in Supplementary Table S4. Pairwise improvements were significant by DeLong’s test (**P<0.001**; Fig. 4a).

**Figure 6.**
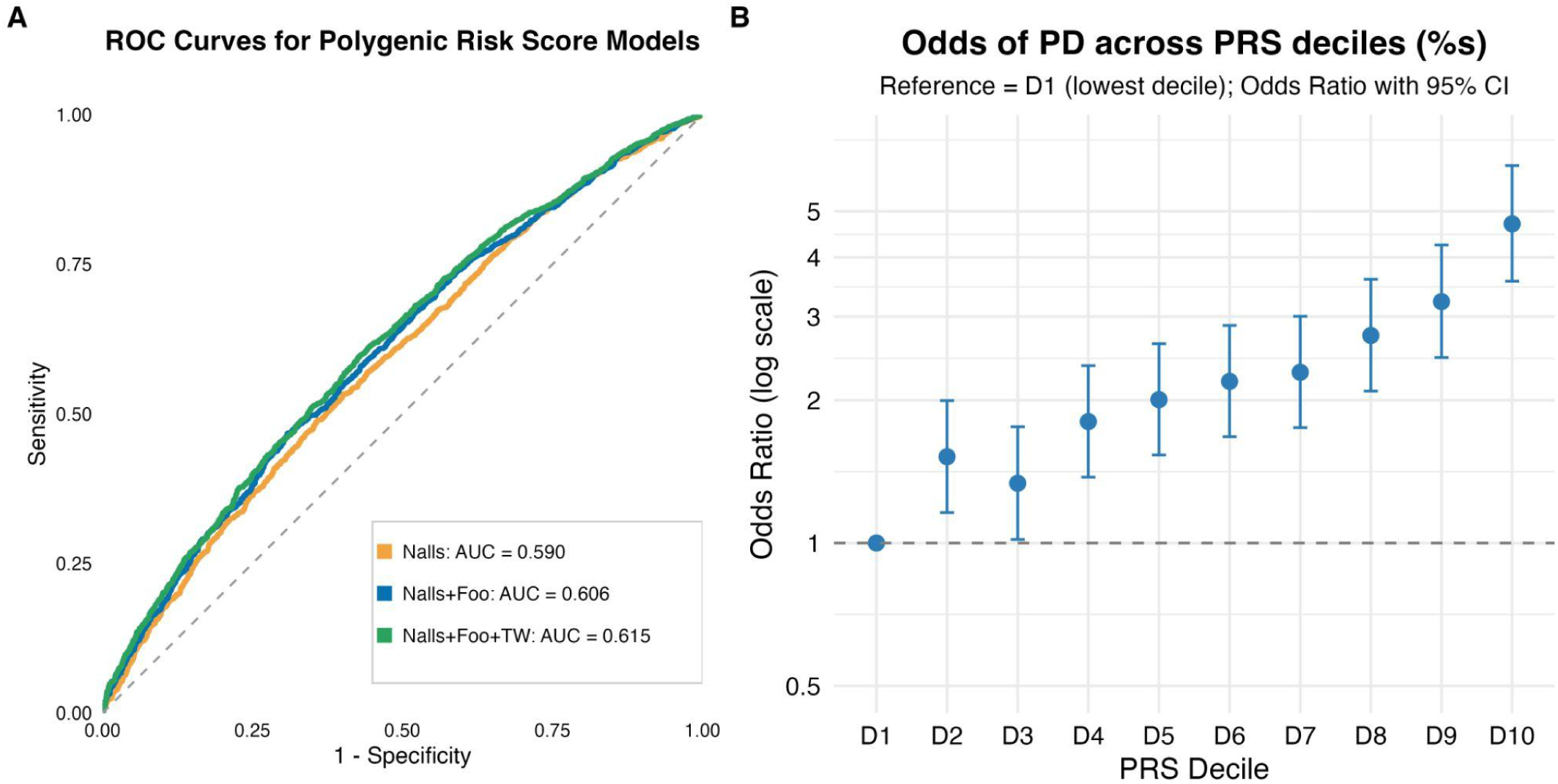
Polygenic risk score (PRS) performance and stratified risk of Parkinson’s disease in the Taiwanese cohort. **(A)** Receiver operating characteristic (ROC) curves comparing three PRS models: **PRS-EU** (variants from Nalls et al., 2019), **PRS-Asia** (PRS-EU plus independent Asian variants from Foo et al., 2020; overlapping loci removed), and **PRS-TW** (PRS-Asia plus Taiwan-specific variants: *LRRK2* p.G2385R, *LRRK2* p.R1628P, *VPS13C* rs12900645, and *SCARB2/FAM47E* rs76591264). Discrimination improved stepwise from **AUC 0.590** to **0.606** to **0.615**; each increment was significant (DeLong’s test, **p < 0.001**). SNP composition and effect weights are listed in **Supplementary Table 3**. **(B)** PRS decile–based risk stratification. Participants were grouped into ten PRS deciles with D1 as reference. Parkinson’s disease risk rose monotonically across deciles, reaching **OR 5.46** (95% CI, 4.09–7.29) in D10 versus D1. Odds ratios and 95% CIs were estimated by logistic regression adjusted for age, sex, and the top ten ancestry principal components. Exact values for all deciles are provided in **Supplementary Table 4**.

Risk stratification by PRS deciles demonstrated a clear dose–response: odds ratios increased monotonically from D2 (OR=**1.52**, 95% CI 1.16–2.00) to D10 (OR=**5.46**, 95% CI 4.09–7.29) relative to D1, adjusted for age, sex, and ancestry PCs (Fig. 4b; Supplementary Table S5). These findings indicate that incorporating East Asian and Taiwan-specific signals measurably enhances PD risk discrimination in this population.

## Discussion

This is the first large-scale genome-wide association study (GWAS) of Parkinson’s disease (PD) in a Taiwanese cohort. It refines PD genetics in East Asia and underscores the value of ancestry-specific analyses. We replicated core PD loci (*SNCA, MCCC1, GCH1*) and East Asian *LRRK2* risk alleles (p.G2385R and p.R1628P), supporting conserved biological pathways across ancestries.^2,3,14^ At the same time, we observed clear ancestry-patterned features at *SNCA*. The Taiwanese lead variants **rs3775427** and **rs5860181** (both intron 4) reside on an East Asian–enriched haplotype. LDlink analyses using 1000 Genomes East Asian data showed tight correlation with **rs6826785** (D′≈0.996, r²≈0.988) but only partial correlation with the European lead **rs356182** (D′≈0.83, r²≈0.47), consistent with pan-Asian GWAS architecture.^2,3,9^ Conditional analyses reinforced this pattern: the intron-4 variants **rs3775437** and **rs5860181** remained associated when conditioning on **rs356182**, whereas the effect at **rs356182** was attenuated when conditioning on **rs5860181**. Notably, the LARGE-PD consortium, which performed a similar conditional analysis (conditioning on rs356182), did not observe any residual intron-4 signal^15^. This contrast implies that the intron-4 association may be more prominent in the Taiwanese and broader East Asian populations.

Haplotype-based association identified two protective configurations (**AGTTG**, **AATTG**) that both carry rs3775427-G; notably, **AGTTG** retains rs356182-G on the canonical East Asian background. Together, these findings indicate that variation within the intron 4 region contributes meaningfully to disease risk, and that SNCA risk and protection in Taiwan are better understood in the context of regional haplotypes rather than single variants. This has translational relevance as α-synuclein–targeted assays and therapies evolve, since underlying genetic polymorphisms may differentially influence α-synuclein expression, conformation, and treatment response.^16^

In *LRRK2*, both p.R1628P and p.G2385R conferred risk in line with prior East Asian studies,^5^ whereas the commonly reported East Asian “protective” combination p.N551K–p.R1398H was not significantly protective in this cohort^17^. The combined carrier frequency of p.R1628P and p.G2385R was 10.8% in controls and 16.3% in PD (Table 4), indicating that these alleles contribute substantially to population-level susceptibility. At the population level, p.R1628P and p.G2385R in *LRRK2* reside on distinct, largely independent haplotypes. Projection of study genotypes onto the 1000 Genomes East Asian subpopulations (CHB, JPT, CHS, CDX, KHV) showed that p.G2385R is present in CHB and JPT, consistent with prior reports in Japanese and Korean cohorts ^18,19^.A founder-effect analysis has further suggested that p.G2385R carriers share a most recent common ancestor ∼4.8 kya^20^. By contrast, p.R1628P is observed predominantly in CHS, CDX (Chinese Dai population in Xishuangbanna), and KHV (Kinh in Ho Chi Minh City, Vietnam), indicating a more southern distribution; its origin remains unclear. These contrasting geographic patterns underscore heterogeneous ancestral backgrounds in the Taiwanese cohort and highlight the differential population structure of East Asian *LRRK2* risk variants.

Genotype-burden analyses supported a dose–response: **single-variant carriers** had **OR = 1.49** (95% CI 1.23–1.79, **P = 4.1 × 10**⁻⁵) versus noncarriers, while **double-variant carriers** (p.G2385R/p.G2385R, p.R1628P/p.R1628P, or compound p.G2385R/p.R1628P) had **OR = 4.30** (95% CI 1.59–11.6, **P = 0.004**) versus noncarriers and **OR = 2.89** (95% CI 1.06–7.91, **P = 0.039**) versus single-variant carriers, consistent with an additive gene-dosage effect of East Asian *LRRK2* alleles. Compound heterozygotes (p.G2385R/p.R1628P) showed a non-significant trend toward earlier age at onset (**55.6 ± 5.8** vs **60.8 ± 11.2** years in noncarriers; t-test **P = 0.06**), suggesting potential synergistic effect between these 2 variants.^21^

Given the relative enrichment of double-variant carriers (**0.8%** of PD cases vs **0.2%** of controls), these individuals represent a priority group for prodromal PD research and preventive intervention. Notably, caffeine intake may modify PD risk among *LRRK2* variant carriers in Asian populations^22^, and first-in-class **type II kinase inhibitors** that stabilize the inactive *LRRK2* conformation have recently been reported^23^, highlighting opportunities for pharmacogenomic stratification and targeted prevention.

At *GCH1*, we replicated association at **rs3783640**, which is in LD with the previously reported **rs11158026** signal^24^; in contrast, **rs841-A** alone was not protective in our data, and haplotype analysis suggested that earlier protective observations attributed to rs841 are likely mediated by the correlation with rs11158026-T^25^. For *MCCC1*, the Taiwanese effect size (OR≈1.32) exceeded European reports (≈1.1) and aligns with observations in southern Chinese cohorts.^26^ We also observed a suggestive association at ***PPARGC1A***, a master regulator of mitochondrial biogenesis/oxidative stress previously linked to PD risk^27^ or age at onset^28^ and to essential tremor (ET)^29^; rare-variant data in Han Chinese further support involvement of ***PPARGC1A*** in PD^27^, motivating cross-disorder analyses in Taiwan. ***PPARGC1A*** is a biologically plausible therapeutic node (PGC-1α). In Taiwan’s National Health Insurance Database, exposure to the PPARγ agonist **pioglitazone** was associated with a lower incidence of PD (adjusted HR **0.66**, 95% CI **0.57–0.78**; dose-dependent)^30^, whereas a phase 2 trial in early PD did not demonstrate disease-modifying effects.^31^ These apparently discordant findings suggest context-dependent efficacy. We hypothesize that the pioglitazone–PD association is modified by *PPARGC1A* genotype, warranting genotype-stratified pharmacoepidemiology and prospective validation studies.

Finally, signals within ***VPS13C*** and ***SCARB2*** not in high LD with previously reported leads suggest potential Taiwan-specific haplotypes that warrant replication and conditional fine-mapping^24^. In ***VPS13C***, we detected an intronic protective association at rs12900645 that lies within a previously described protective region (chr15:61,920,582–62,023,836) encompassing the p.R153H–p.I398I–p.I1132V–p.Q2376Q haplotype reported by Rudakou et al.^13^. Linkage disequilibrium between rs12900645 and p.I1132V was modest (r²≈0.44), and the four coding/synonymous variants showed protective but non-significant effects in our data, together suggesting a shared protective background with rs12900645 as the lead tag in Taiwanese individuals. Given that VPS13C encodes a lipid-transfer protein implicated in lysosomal–mitochondrial homeostasis, this signal is biologically plausible but requires confirmation. Replication in independent East Asian cohorts, conditional fine-mapping to establish independence from nearby variants, and functional follow-up (eQTL/splicing colocalization in brain, enhancer perturbation, and lipid-trafficking assays) will be critical to validate a causal mechanism for protection at this locus.

***GALNT13***, a glycosyltransferase, emerged as a putative novel locus, echoing prior Asian evidence at ***WBSCR17*/*GALNT17***^3^ and raising the possibility that O-glycosylation pathways contribute to PD susceptibility in Asian populations^32^. Recent studies have also shown that glycation of α-synuclein promotes its aggregation and exacerbates neuroinflammatory responses.^33^

Cross-ancestry comparisons showed effect sizes that correlated modestly with European meta-analysis and strongly with pan-East Asian estimates, reflecting both shared biology and ancestry-patterned architecture.^2,3^ Consistent with known limits on PRS portability, a European-derived PRS achieved **AUC 0.590** in our cohort; augmenting with Asian variants increased AUC to **0.606**, and further adding Taiwan-specific variants yielded **AUC 0.615** (all pairwise DeLong tests **P<0.001**). These stepwise gains echo broader benchmarking that emphasizes the need for ancestry-aware risk modeling in PD^11^. Methodologically, our region-focused haplotype analyses at *SNCA* and *LRRK2* clarified relationships among closely linked variants and exposed protective/risk configurations that single-SNP tests can miss—an approach that can improve locus interpretation and fine-mapping in diverse cohorts^34^.

Several limitations merit mention. First, we used SNP arrays with imputation rather than comprehensive sequencing, which reduces sensitivity to rare coding variants and structural variation and performs suboptimally at complex loci (e.g., *GBA1* with its pseudogene)^35^. Deep whole-genome sequencing and/or long-read approaches with locus-specific validation would address this. Second, this is a Taiwan-focused cohort designed to fill a clear gap; findings may not capture the full heterogeneity across East and Southeast Asia. Replication in independent EAPDGC sites^12^ and additional regional cohorts will be important. Third, PRS performance was assessed using logistic regression without clinical or lifestyle covariates (e.g., caffeine^22^, smoking), prodromal markers, or biomarkers. Future work should evaluate integrative models that combine genetics with environmental and clinical features, consider machine-learning approaches^36^ with rigorous internal/external validation, and report calibration and clinical utility (e.g., decision-curve analysis).^37^

In conclusion, this Taiwan-wide GWAS refines PD genetics in East Asia and underscores the value of ancestry-specific analysis. We confirm core loci (*SNCA, MCCC1, GCH1*), delineate an East Asian intron-4 *SNCA* haplotype driving risk/protection, and show that *LRRK2* p.G2385R/p.R1628P are common with a clear gene-dosage effect—prioritizing double-variant carriers for prodromal and prevention studies. We also highlight a protective region at *VPS13C* and suggestive signals at *PPARGC1A* and *GALNT13*, pointing to mitochondrial, lysosomal/trafficking, and glycosylation pathways. A Taiwan-optimized PRS outperforms a European model (AUC 0.590→0.615). Replication across East/Southeast Asia, fine-mapping, functional validation, and integration with clinical/environmental data are warranted.

## Methods

### Study population and cohort design

Participants were recruited through the East Asian Parkinson Disease Genomics Consortium (EAPDGC) via a multi-center collaboration. Taiwanese recruitment sites included National Taiwan University Hospital (NTUH), Chang Gung Memorial Hospital (CGMH), Shuang Ho Hospital/Taipei Medical University (SHH/TMU), Landseed Hospital and Taiwan Biobank.^38,39^ Parkinson’s disease (PD) diagnosis followed the Movement Disorder Society (MDS) Clinical Diagnostic Criteria.^40^ All participants provided written informed consent. The study protocol was approved by the NTUH Institutional Review Board (NTUH-REC No. 201905035RINC).

### Genotyping and quality control

Genotyping used the Illumina NeuroBooster Array, which combines the Global Diversity Array backbone with custom neuro-focused content.^41^ Quality control (QC) was performed in PLINK v1.9 and v2.0.^42,43^ Samples were excluded for low call rate (<95%), sex discordance, or excess relatedness (one of each pair with PI_HAT >0.20 removed). Variants were removed for Hardy–Weinberg disequilibrium in controls (p<1×10⁻⁴), missingness >5%, or minor allele frequency (MAF) <0.005. Principal component analysis (PCA) was performed to control for population structure. Association models adjusted for age, sex, and ancestry PCs derived from study genotypes.

### Imputation

Post-QC genotypes were imputed using the Taiwan Biobank (TWB) imputation service, which employs Minimac4 and a population-specific reference panel of ∼1,500 TWB whole-genome sequences.^38,44^ Imputation was performed on the TWB server pipeline; we applied post-imputation filters of MAF>0.01 and Rsq>0.30, yielding ∼7.6 million high-quality variants for analysis.

### Principal Component Analysis (PCA)

Autosomal, biallelic SNPs were LD-pruned in PLINK v1.9 (parameters: --maf 0.10; --indep 50 5 1.5), and principal components were then computed on the pruned set using --pca. Study genotypes were merged with the 1000 Genomes Project Phase 3 (GRCh38) reference to anchor ancestry inference. Scatter plots of top PC were generated in R (v4.4.2) using ggplot2 to visualize clustering; participants aligned with the East Asian superpopulation (Figure 3 and Supplementary Fig. S1).

### Statistical analysis

Genome-wide association testing used logistic regression in PLINK 2.0 with PD status as the outcome, adjusting for age, sex, and the top 15 principal components. Genome-wide significance was defined as p<5×10^-8^ and suggestive significance as p<1×10^-5^. Analyses were later conducted in R (v4.4.2).^45^ Specifically, we ran logistic regression in PLINK 2.0 using the --condition flag to include the index variant as a covariate, adjusting for age, sex, and the first 15 principal components.

### Linkage disequilibrium and haplotype analysis

Population LD context was evaluated using LDlink to query East Asian reference panels.¹² Within the Taiwanese cohort, pairwise LD (r²) was estimated with PLINK (–r2). Haplotype visualization for *SNCA*, *LRRK2*, and *GCH1* used the **geneHapR** R package^46^; haplotype association testing employed **haplo.stats** ^47^with EM-based haplotype frequency estimation, adjusting for age, sex, and the first five PCs.

### Cross-ancestry replication and effect-size correlation

To assess cross-population consistency, we extracted significant variants from the European meta-analysis (Nalls et al., 2019) and the pan-Asian GWAS (Foo et al., 2020), harmonized alleles to GRCh38, and computed Pearson correlations of log-odds ratios between this study and reference datasets.

### Polygenic risk score (PRS) construction and evaluation

We constructed three PRS models: (i) **PRS-EU**, based on variants reported in the Nalls et al. 2019 European meta-analysis; (ii) **PRS-Asia**, augmenting PRS-EU with independent risk variants from Foo et al. 2020; and (iii) **PRS-TW**, further augmenting PRS-Asia with variants identified as significant or suggestive in the Taiwanese GWAS. Model performance was evaluated by logistic regression (adjusted for age, sex, and PCs) and compared using area under the receiver operating characteristic curve (AUC) with DeLong’s test for paired ROC curves.^48^

## Acknowledgements

This work was conducted as part of the East Asian Parkinson Disease Genomics Consortium (EAPDGC). The EAPDGC is supported by the Michael J. Fox Foundation for Parkinson’s Research through the Genetic Diversity in Parkinson’s Disease initiative (Grant No. 17474). This project was supported by the Global Parkinson’s Genetics Program (GP2; https://gp2.org). GP2 is funded by the Aligning Science Across Parkinson’s (ASAP) initiative and implemented by The Michael J. Fox Foundation for Parkinson’s Research (MJFF). For a complete list of GP2 members see https://doi.org/10.5281/zenodo.7904831. For the purpose of Open Access, the author has applied a CC-BY public copyright licence to any Author Accepted Manuscript version arising from this submission, the Global Parkinson’s Genetics Program (GP2), and Aligning Science Across Parkinson’s (ASAP). We also thank the Taiwan Biobank for providing control samples and support with genotype imputation, and Prof. Andrew B. Singleton and Prof. Matthew J. Farrer for their constructive comments on the analyses and manuscript.

## Data Availability

Summary statistics generated in this study will be made available through the GP2 platform. Access to the individual-level data used in the preparation of this article is coordinated by the corresponding authors and governed by the GP2 Tier 2 data access policy, and may be granted upon approval of the request and execution of a Data Use Agreement by the applicant’s institution. GP2 data are available via the AMP PD platform (https://amp-pd.org). Summary statistics from Nalls et al. were retrieved from the GWAS Catalog (https://www.ebi.ac.uk/gwas/), and summary statistics from Foo et al. were obtained from the supplementary materials of the original publication in JAMA Neurology.^3^

## Code Availability

All code generated for this article, and the identifiers for all software programs and packages used, are available on GitHub https://github.com/GP2code/Taiwan-GWAS-and-PRS and were given a persistent identifier via Zenodo DOI 10.5281/zenodo.18232864.

## Author Contributions

Y.T.C. collected clinical data, performed statistical analyses, and drafted the manuscript. K.Y.M. and R.M.W. conceived and designed the study. Y.A.S. performed genotyping. C.H.L., C.H.T., Y.R.W., C.T.H., and Y.W.C. recruited participants and acquired clinical data. M.H.T. supervised genotyping and data analysis. J.H. provided resources and critical intellectual input. All authors reviewed and approved the final manuscript.

## References

1. Bloem, B. R., Okun, M. S. & Klein, C. Parkinson’s disease. Lancet 397, 2284–2303 (2021).

2. Nalls, M. A. et al. Identification of novel risk loci, causal insights, and heritable risk for Parkinson’s disease: a meta-analysis of genome-wide association studies. Lancet Neurol 18, 1091–1102 (2019).

3. Foo, J. N. et al. Identification of Risk Loci for Parkinson Disease in Asians and Comparison of Risk Between Asians and Europeans A Genome-Wide Association Study. Jama Neurol. 77, 746–754 (2020).

4. Lim, S.-Y. et al. Parkinson’s disease in the Western Pacific Region. Lancet Neurol. 18, 865–879 (2019).

5. Ross, O. A. et al. Association of LRRK2 exonic variants with susceptibility to Parkinson’s disease: a case-control study. Lancet Neurol. 10, 898–908 (2011).

6. Tan, E. K. et al. Multiple LRRK2 variants modulate risk of Parkinson disease: a Chinese multicenter study. Hum Mutat 31, 561–8 (2010).

7. Zhao, H. & Kong, Z. Relationship between LRRK2 R1628P polymorphism and Parkinson’s disease in Asian populations. Oncotarget 7, 46890–46898 (2016).

8. Loesch, D. P. et al. Polygenic risk prediction and SNCA haplotype analysis in a Latino Parkinson’s disease cohort. Parkinsonism Relat. Disord. 102, 7–15 (2022).

9. Pihlstrøm, L. et al. A comprehensive analysis of SNCA-related genetic risk in sporadic parkinson disease. Ann. Neurol. 84, 117–129 (2018).

10. Rizig, M. et al. Identification of genetic risk loci and causal insights associated with Parkinson’s disease in African and African admixed populations: a genome-wide association study. Lancet Neurol. 22, 1015–1025 (2023).

11. Saffie-Awad, P. et al. Insights into ancestral diversity in Parkinson’s disease risk: a comparative assessment of polygenic risk scores. NPJ Park. Dis. 11, 201 (2025).

12. Mok, K. Y. The East Asian Parkinson Disease Genomics Consortium. Lancet Neurol 20, 982 (2021).

13. Rudakou, U. et al. Analysis of common and rare VPS13C variants in late-onset Parkinson disease. Neurol. Genet. 6, 385 (2020).

14. Bandres-Ciga, S. et al. NeuroBooster Array: A Genome-Wide Genotyping Platform to Study Neurological Disorders Across Diverse Populations. medRxiv 10.1101/2023.11.06.23298176 (2023) doi:10.1101/2023.11.06.23298176.

15. Loesch, D. P. et al. Characterizing the Genetic Architecture of Parkinson’s Disease in Latinos. Ann. Neurol. 90, 353–365 (2021).

16. Li, D., Yau, W. Y., Chen, S., Wilton, S. & Mastaglia, F. A personalised and comprehensive approach is required to suppress or replenish SNCA for Parkinson’s disease. NPJ Park. Dis. 11, 42 (2025).

17. Gopalai, A. A., et al. LRRK2 N551K and R1398H variants are protective in Malays and Chinese in Malaysia: A case-control association study for Parkinson’s disease. Mol. Genet. Genomic Med. 7, e604 (2019).

18. Zabetian, C. P. et al. LRRK2 Mutations and Risk Variants in Japanese Patients with Parkinson’s Disease. Mov. Disord. Off. J. Mov. Disord. Soc. 24, 1034–1041 (2009).

19. Kim, J.-M. et al. The LRRK2 G2385R variant is a risk factor for sporadic Parkinson’s disease in the Korean population. Parkinsonism Relat. Disord. 16, 85–88 (2010).

20. Farrer, M. J. et al. Lrrk2 G2385R is an ancestral risk factor for Parkinson’s disease in Asia. Parkinsonism Relat. Disord. 13, 89–92 (2007).

21. Goh, J. W. et al. LRRK2 p.G2385R and p.R1628P Variants in a Multi-Ethnic Asian Parkinson’s Cohort: Epidemiology and Clinical Insights. 2025.08.06.25332985 Preprint at 10.1101/2025.08.06.25332985 (2025).

22. Ong, Y.-L. et al. Caffeine intake interacts with Asian gene variants in Parkinson’s disease: a study in 4488 subjects. Lancet Reg. Health West. Pac. 40, 100877 (2023).

23. Type II kinase inhibitors that target Parkinson’s disease–associated LRRK2 | Science Advances. https://www.science.org/doi/10.1126/sciadv.adt2050.

24. Chen, C.-M. et al. Association of GCH1 and MIR4697, but not SIPA1L2 and VPS13C polymorphisms, with Parkinson’s disease in Taiwan. Neurobiol. Aging 39, 221.e1–5 (2016).

25. Rudakou, U. et al. Common and rare GCH1 variants are associated with Parkinson disease. Neurobiol. Aging 73, 231.e1–231.e6 (2019).

26. Zhao, A. et al. SNPs in SNCA, MCCC1, DLG2, GBF1 and MBNL2 are associated with Parkinson’s disease in southern Chinese population. J. Cell. Mol. Med. 24, 8744–8752 (2020).

27. Li, L.-Z. et al. Association of rare PPARGC1A variants with Parkinson’s disease risk. J. Hum. Genet. 67, 687–690 (2022).

28. Clark, J., Reddy, S., Zheng, K., Betensky, R. A. & Simon, D. K. Association of PGC-1alpha polymorphisms with age of onset and risk of Parkinson’s disease. BMC Med. Genet. 12, 69 (2011).

29. Müller, S. H. et al. Genome-wide association study in essential tremor identifies three new loci. Brain J. Neurol. 139, 3163–3169 (2016).

30. Chen, L., Tao, Y., Li, J. & Kang, M. Pioglitazone use is associated with reduced risk of Parkinson’s disease in patients with diabetes: A systematic review and meta-analysis. J. Clin. Neurosci. 106, 154–158 (2022).

31. NINDS Exploratory Trials in Parkinson Disease (NET-PD) FS-ZONE Investigators. Pioglitazone in early Parkinson’s disease: a phase 2, multicentre, double-blind, randomised trial. Lancet Neurol. 14, 795–803 (2015).

32. Wenzel, D. M. & Olivier-Van Stichelen, S. The O-GlcNAc cycling in neurodevelopment and associated diseases. Biochem. Soc. Trans. 50, 1693–1702 (2022).

33. Vasili, E. et al. Glycation of alpha-synuclein enhances aggregation and neuroinflammatory responses. Npj Park. Dis. 11, 307 (2025).

34. Schaid, D. J., Rowland, C. M., Tines, D. E., Jacobson, R. M. & Poland, G. A. Score tests for association between traits and haplotypes when linkage phase is ambiguous. Am. J. Hum. Genet. 70, 425–434 (2002).

35. Pachchek, S. et al. Accurate long-read sequencing identified GBA1 as major risk factor in the Luxembourgish Parkinson’s study. NPJ Park. Dis 9, 156 (2023).

36. Makarious, M. B. et al. Multi-modality machine learning predicting Parkinson’s disease. Npj Park. Dis. 8, 35 (2022).

37. Kullo, I. J. Clinical use of polygenic risk scores: current status, barriers and future directions. Nat. Rev. Genet. 10.1038/s41576-025-00900-8 (2025) doi:10.1038/s41576-025-00900-8.

38. Feng, Y.-C. A. et al. Taiwan Biobank: A rich biomedical research database of the Taiwanese population. Cell Genomics 2, 100197 (2022).

39. Chen, C.-H. et al. Population structure of Han Chinese in the modern Taiwanese population based on 10,000 participants in the Taiwan Biobank project. Hum. Mol. Genet. 25, 5321–5331 (2016).

40. Postuma, R. B. et al. MDS clinical diagnostic criteria for Parkinson’s disease. Mov Disord 30, 1591–601 (2015).

41. Bandres-Ciga, S. et al. NeuroBooster Array: A Genome-Wide Genotyping Platform to Study Neurological Disorders Across Diverse Populations. Mov. Disord. Off. J. Mov. Disord. Soc. 39, 2039–2048 (2024).

42. Purcell, S. et al. PLINK: A Tool Set for Whole-Genome Association and Population-Based Linkage Analyses. Am. J. Hum. Genet. 81, 559–575 (2007).

43. Chang, C. C. et al. Second-generation PLINK: rising to the challenge of larger and richer datasets. GigaScience 4, 7 (2015).

44. Das, S. et al. Next-generation genotype imputation service and methods. Nat Genet 48, 1284–1287 (2016).

45. R: a language and environment for statistical computing. https://www.gbif.org/tool/81287/r-a-language-and-environment-for-statistical-computing.

46. Zhang, R., Jia, G. & Diao, X. geneHapR: an R package for gene haplotypic statistics and visualization. BMC Bioinformatics 24, 199 (2023).

47. Daniel, S. & Sinnwell, J. P. haplo.stats: Statistical Analysis of Haplotypes with Traits and Covariates when Linkage Phase is Ambiguous. (2024).

48. DeLong, E. R., DeLong, D. M. & Clarke-Pearson, D. L. Comparing the areas under two or more correlated receiver operating characteristic curves: a nonparametric approach. Biometrics 44, 837–845 (1988).

