## Supplementary material for "Genome-Wide Association and Population-Tailored Polygenic Risk for Parkinson’s Disease in Taiwan": Supplmental Table and Figure

**Supplementary Table S1. Recruitment centers and sample sizes**
List of hospitals and institutions contributing PD cases and controls

| | Case before QC | Case After QC | Control Before QC | Control After QC |
|---|---|---|---|---|
| NTUH | 1916 | 1850 | 900 | 846 |
| CGMH | 317 | 290 | 484 | 451 |
| Taiwan Biobank | | | 530 | 476 |
| SHH and TMU Biobank | 166 | 111 | 400 | 396 |
| Landseed Biobank | | | 130 | 99 |
| Total Number | 2399 | 2251 | 2444 | 2268 |

**Supplementary Table S2: Full Genetic Risk Loci Associated with PD in Taiwan** with p value less than 1e-5

| Chr | Position (hg38) | SNP (rsID) | Nearest Gene | Effect Allele | Other Allele | MAF | OR_CI | β | SE | P | Annotation |
|---|---|---|---|---|---|---|---|---|---|---|---|
| 2 | 154088802 | rs10186542 | *GALNT13* | A | G | 0.12 | 1.35 (1.18–1.55) | 0.13 | 0.07 | 5.30e-06 | New |
| 2 | 179727418 | rs4894119 | *ZNF385B* | G | A | 0.02 | 2.11 (1.54–2.89) | 0.32 | 0.16 | 5.20e-06 | New |
| 3 | 183017423 | rs2292056 | *MCCC1* | T | G | 0.43 | 1.31 (1.21–1.42) | 0.12 | 0.04 | 8.50e-10 | Foo |
| 3 | 183028420 | rs34822404 | *MCCC1* | C | T | 0.43 | 1.31 (1.21–1.42) | 0.12 | 0.04 | 7.20e-10 | Most significat in Taiwan |
| 3 | 183042285 | rs10513789 | *MCCC1* | T | G | 0.43 | 1.3 (1.20–1.41) | 0.12 | 0.04 | 1.40e-09 | Nalls |
| 4 | 24009221 | rs609501 | *PPARGC1A* | A | G | 0.48 | 1.22 (1.13–1.32) | 0.09 | 0.04 | 3.50e-06 | New |
| 4 | 89704960 | rs356182 | *SNCA* | A | G | 0.3 | 0.78 (0.71–0.86) | -0.11 | 0.05 | 1.90e-07 | Nalls |
| 4 | 89744890 | rs356203 | *SNCA* | T | C | 0.44 | 0.77 (0.71–0.83) | -0.11 | 0.04 | 9.50e-10 | Past European |
| 4 | 89759478 | rs5860181 | *SNCA* | A | ATGC ATATT | 0.43 | 0.75 (0.69–0.81) | -0.12 | 0.04 | 3.70e-11 | Most significat in Taiwan |
| 4 | 89761323 | rs6826785 | *SNCA* | T | C | 0.43 | 0.76 (0.70–0.82) | -0.12 | 0.04 | 1.60e-10 | Foo |
| 4 | 89765968 | rs3775427 | *SNCA* | G | A | 0.43 | 0.75 (0.69–0.81) | -0.12 | 0.04 | 6.90e-11 | Most significat in Taiwan |
| 5 | 119707094 | rs2194056 | *AC008550.1* | C | G | 0.04 | 0.57 (0.45–0.72) | -0.24 | 0.12 | 1.60e-06 | New |
| 5 | 131320353 | rs185601781 | *CDC42SE2* | G | C | 0.02 | 0.52 (0.39–0.70) | -0.29 | 0.15 | 5.40e-06 | New |
| 7 | 153256105 | rs12671235 | *LOC102723686* | A | T | 0.32 | 0.8 (0.73–0.88) | -0.09 | 0.05 | 2.70e-06 | New |
| 9 | 102260653 | rs76204651 | *RNU6-329P* | A | G | 0.06 | 0.65 (0.54–0.78) | -0.18 | 0.09 | 5.80e-06 | New |
| 14 | 54885803 | rs3783640 | *GCH1* | T | C | 0.46 | 1.23 (1.14–1.33) | 0.09 | 0.04 | 7.70e-07 | Taiwan |
| 14 | 54882151 | rs11158026 | *GCH1* | C | T | 0.46 | 1.23 (1.14–1.33) | 0.09 | 0.04 | 1.40e-06 | Nalls |
| 17 | 43331428 | rs78543719 | *CCDC200* | A | G | 0.02 | 0.45 (0.32–0.63) | -0.35 | 0.17 | 1.70e-06 | New |
| 17 | 70673898 | rs17792098 | *AC005771.1* | G | A | 0.19 | 1.31 (1.16–1.47) | 0.12 | 0.06 | 1.30e-06 | New |
| 18 | 51753748 | rs3844136 | *AC008550.1* | G | A | 0.06 | 0.65 (0.54–0.78) | -0.19 | 0.09 | 4.30e-06 | New |

**Supplementary Table S3. Comparison of known PD loci across European, Asian, and Taiwanese GWAS**

Effect size and p-value from Nalls et al. (2019, European), Foo et al. (2020, Asian), and current Taiwanese GWAS.

| source | rsID | Nearest Gene | chr | position hg38 | A1 | REF | A1_FREQ | OR | logOR | P | EAS Frequency | EUR Frequency | Nalls effect weight/ logOR |
|---|---|---|---|---|---|---|---|---|---|---|---|---|---|
| Nalls 2019 | rs114138760 | *PMVK* | 1 | 154925709 | NA | NA | NA | NA | NA | NA | None | 0.01 | 0.28 |
| Nalls 2019 | rs35749011 | *None* | 1 | 155162560 | NA | NA | NA | NA | NA | NA | None | 0.02 | 0.61 |
| Nalls 2019 | rs76763715 | *GBA* | 1 | 155235843 | NA | NA | NA | NA | NA | NA | None | 0.00 | -0.75 |
| Nalls 2019 | rs6658353 | *None* | 1 | 161499264 | C | G | 0.30 | 1.05 | 0.05 | 0.33 | 0.22 | 0.51 | 0.07 |
| Nalls 2019 | rs11578699 | *None* | 1 | 171750629 | T | C | 0.18 | 0.96 | -0.04 | 0.47 | 0.16 | 0.17 | -0.07 |
| Nalls 2019 | rs823118 | *None* | 1 | 205754444 | C | T | 0.40 | 0.86 | -0.15 | 0.00 | 0.51 | 0.53 | 0.11 |
| Nalls 2019 | rs11557080 | *RAB29* | 1 | 205768611 | G | A | 0.40 | 0.86 | -0.16 | 0.00 | 0.51 | 0.13 | 0.13 |
| Nalls 2019 | rs4653767 | *ITPKB* | 1 | 226728377 | C | T | 0.27 | 0.90 | -0.10 | 0.03 | 0.26 | 0.31 | 0.08 |
| Nalls 2019 | rs10797576 | *SIPA1L2* | 1 | 232528865 | T | C | 0.14 | 1.02 | 0.02 | 0.72 | 0.15 | 0.14 | 0.11 |
| Nalls 2019 | rs76116224 | *KCNS3* | 2 | 17966582 | NA | NA | NA | NA | NA | NA | 0.00 | 0.09 | 0.11 |
| Nalls 2019 | rs2042477 | *KCNIP3* | 2 | 95335195 | A | T | 0.44 | 1.05 | 0.04 | 0.30 | 0.59 | 0.76 | -0.07 |
| Nalls 2019 | rs11683001 | *MAP4K4* | 2 | 101780501 | A | T | 0.48 | 1.03 | 0.03 | 0.56 | 0.49 | 0.35 | 0.07 |
| Nalls 2019 | rs57891859 | *TMEM163* | 2 | 134707046 | A | G | 0.25 | 1.02 | 0.02 | 0.70 | 0.76 | 0.31 | 0.08 |
| Nalls 2019 | rs1474055 | *None* | 2 | 168253884 | T | C | 0.34 | 1.05 | 0.05 | 0.30 | 0.33 | 0.12 | 0.18 |
| Nalls 2019 | rs73038319 | *TBC1D5* | 3 | 18320267 | NA | NA | NA | NA | NA | NA | None | 0.03 | -0.17 |

| | | | | | | | | | | | | | |
|---|---|---|---|---|---|---|---|---|---|---|---|---|---|
| Nalls 2019 | rs6808178 | *AC098650.1* | 3 | 28664199 | T | C | 0.20 | 1.11 | 0.10 | 0.06 | 0.78 | 0.63 | 0.07 |
| Nalls 2019 | rs6808178 | *RBMS3* | 3 | 28664199 | T | C | 0.20 | 1.11 | 0.10 | 0.06 | 0.78 | 0.63 | 0.07 |
| Nalls 2019 | rs12497850 | *IP6K2* | 3 | 48711556 | G | T | 0.04 | 0.90 | -0.10 | 0.37 | 0.95 | 0.66 | 0.06 |
| Nalls 2019 | rs55961674 | *KPNA1* | 3 | 122478045 | T | C | 0.13 | 0.99 | -0.01 | 0.86 | 0.11 | 0.17 | 0.09 |
| Nalls 2019 | rs11707416 | *MED12L* | 3 | 151391177 | A | T | 0.14 | 1.01 | 0.01 | 0.83 | 0.14 | 0.38 | -0.06 |
| Nalls 2019 | rs1450522 | *SPTSSB* | 3 | 161359842 | A | G | 0.44 | 1.01 | 0.01 | 0.86 | 0.54 | 0.31 | -0.06 |
| Nalls 2019 | rs10513789 | *MCCC1* | 3 | 183042285 | T | G | 0.43 | 1.30 | 0.27 | 0.00 | 0.57 | 0.20 | 0.15 |
| Nalls 2019 | rs873786 | *GAK* | 4 | 931588 | NA | NA | NA | NA | NA | NA | 0.00 | 0.10 | -0.17 |
| Nalls 2019 | rs34311866 | *TMEM175* | 4 | 958159 | C | T | 0.13 | 1.16 | 0.15 | 0.02 | 0.13 | 0.19 | -0.21 |
| Nalls 2019 | rs4698412 | *BST1* | 4 | 15735725 | A | G | 0.40 | 1.14 | 0.13 | 0.00 | 0.38 | 0.56 | 0.10 |
| Nalls 2019 | rs34025766 | *LCORL* | 4 | 17967188 | A | T | 0.13 | 1.05 | 0.05 | 0.40 | 0.16 | 0.13 | -0.08 |
| Nalls 2019 | rs6825004 | *SCARB2* | 4 | 76189212 | G | C | 0.39 | 0.97 | -0.03 | 0.54 | 0.36 | 0.31 | 0.06 |
| Nalls 2019 | rs4101061 | *SCARB2/ FAM47E* | 4 | 76226816 | A | G | 0.23 | 1.05 | 0.05 | 0.31 | 0.78 | 0.28 | -0.09 |
| Nalls 2019 | rs6854006 | *FAM47E-STBD1* | 4 | 76276901 | T | C | 0.09 | 1.17 | 0.16 | 0.03 | 0.08 | 0.39 | -0.09 |
| Nalls 2019 | rs356182 | *SNCA* | 4 | 89704960 | A | G | 0.30 | 0.78 | -0.24 | 0.00 | 0.37 | 0.64 | -0.28 |
| Nalls 2019 | rs5019538 | *SNCA* | 4 | 89715479 | NA | NA | NA | NA | NA | NA | 1.00 | 0.68 | -0.16 |
| Nalls 2019 | rs13117519 | *None* | 4 | 113447909 | T | C | 0.09 | 1.15 | 0.14 | 0.06 | 0.08 | 0.18 | 0.09 |
| Nalls 2019 | rs62333164 | *NA* | 4 | 169662006 | A | G | 0.06 | 0.83 | -0.18 | 0.04 | NA | NA | -0.06 |
| Nalls 2019 | rs1867598 | *ELOVL7* | 5 | 60842132 | G | A | 0.06 | 1.11 | 0.10 | 0.26 | 0.05 | 0.11 | -0.16 |

| | | | | | | | | | | | | | |
|---|---|---|---|---|---|---|---|---|---|---|---|---|---|
| Nalls 2019 | rs26431 | *PAM* | 5 | 103030090 | G | C | 0.44 | 0.97 | -0.03 | 0.47 | 0.55 | 0.72 | 0.06 |
| Nalls 2019 | rs11950533 | *AC006077.1* | 5 | 134863415 | A | C | 0.24 | 1.00 | 0.00 | 0.96 | 0.31 | 0.14 | -0.09 |
| Nalls 2019 | rs4140646 | *None* | 6 | 27771022 | A | G | 0.22 | 1.12 | 0.11 | 0.06 | 0.18 | 0.22 | 0.08 |
| Nalls 2019 | rs9261484 | *TRIM40* | 6 | 30140906 | T | C | 0.31 | 1.00 | 0.00 | 1.00 | 0.26 | 0.26 | -0.06 |
| Nalls 2019 | rs112485576 | *NA* | 6 | 32610995 | A | C | 0.08 | 0.99 | -0.01 | 0.96 | NA | NA | -0.17 |
| Nalls 2019 | rs12528068 | *None* | 6 | 71778059 | T | C | 0.08 | 1.12 | 0.11 | 0.17 | 0.09 | 0.26 | 0.07 |
| Nalls 2019 | rs997368 | *None* | 6 | 111922088 | G | A | 0.37 | 0.92 | -0.08 | 0.08 | 0.36 | 0.20 | 0.07 |
| Nalls 2019 | rs75859381 | *None* | 6 | 132889222 | C | T | 0.04 | 1.26 | 0.23 | 0.05 | 0.02 | 0.03 | -0.22 |
| Nalls 2019 | rs199351 | *GPNMB* | 7 | 23260430 | C | A | 0.33 | 0.96 | -0.04 | 0.37 | 0.35 | 0.38 | 0.10 |
| Nalls 2019 | rs76949143 | *AC006001.3/AC008267.3* | 7 | 66544864 | A | T | 0.14 | 0.88 | -0.13 | 0.04 | 0.17 | 0.06 | -0.14 |
| Nalls 2019 | rs1293298 | *CTSB* | 8 | 11854934 | NA | NA | NA | NA | NA | NA | 0.00 | 0.26 | 0.09 |
| Nalls 2019 | rs620513 | *AC011586.2* | 8 | 16840084 | T | G | 0.42 | 0.90 | -0.11 | 0.01 | 0.42 | 0.29 | -0.09 |
| Nalls 2019 | rs2280104 | *BIN3* | 8 | 22668467 | T | C | 0.17 | 0.95 | -0.05 | 0.35 | 0.80 | 0.63 | 0.06 |
| Nalls 2019 | rs2086641 | *FAM49B* | 8 | 129889663 | C | T | 0.50 | 1.05 | 0.05 | 0.24 | 0.52 | 0.28 | -0.06 |
| Nalls 2019 | rs13294100 | *SH3GL2* | 9 | 17579692 | G | T | 0.47 | 1.08 | 0.08 | 0.07 | 0.48 | 0.64 | -0.09 |
| Nalls 2019 | rs10756907 | *SH3GL2* | 9 | 17727067 | G | A | 0.39 | 1.04 | 0.04 | 0.32 | 0.37 | 0.23 | -0.09 |
| Nalls 2019 | rs6476434 | *UBAP2* | 9 | 34046393 | C | T | 0.16 | 1.13 | 0.12 | 0.04 | 0.83 | 0.75 | -0.06 |
| Nalls 2019 | rs896435 | *ITGA8* | 10 | 15515407 | T | C | 0.44 | 0.97 | -0.03 | 0.47 | 0.43 | 0.71 | 0.07 |
| Nalls 2019 | rs10748818 | *GBF1* | 10 | 102255522 | G | A | 0.36 | 1.15 | 0.14 | 0.00 | 0.37 | 0.12 | -0.08 |

| | | | | | | | | | | | | | |
|---|---|---|---|---|---|---|---|---|---|---|---|---|---|
| Nalls 2019 | rs72840788 | *BAG3* | 10 | 119656173 | NA | NA | NA | NA | NA | NA | 0.00 | 0.22 | 0.08 |
| Nalls 2019 | rs117896735 | *INPP5F* | 10 | 119776815 | NA | NA | NA | NA | NA | NA | None | 0.01 | 0.44 |
| Nalls 2019 | rs7938782 | *RNF141* | 11 | 10537230 | G | A | 0.16 | 0.93 | -0.07 | 0.22 | 0.14 | 0.15 | 0.09 |
| Nalls 2019 | rs12283611 | *DLG2* | 11 | 83776234 | A | C | 0.25 | 1.07 | 0.06 | 0.20 | 0.28 | 0.43 | -0.06 |
| Nalls 2019 | rs3802920 | *IGSF9B* | 11 | 133917106 | T | G | 0.15 | 0.96 | -0.04 | 0.46 | 0.15 | 0.18 | 0.11 |
| Nalls 2019 | rs76904798 | *LRRK2* | 12 | 40220632 | T | C | 0.05 | 1.09 | 0.08 | 0.39 | 0.06 | 0.13 | 0.14 |
| Nalls 2019 | rs34637584 | *LRRK2* | 12 | 40340400 | NA | NA | NA | NA | NA | NA | None | None | 2.43 |
| Nalls 2019 | rs7134559 | *None* | 12 | 46025303 | C | T | 0.40 | 1.12 | 0.11 | 0.01 | 0.56 | 0.41 | -0.05 |
| Nalls 2019 | rs10847864 | *HIP1R* | 12 | 122842051 | G | T | 0.45 | 0.96 | -0.04 | 0.35 | 0.53 | 0.34 | 0.15 |
| Nalls 2019 | rs11610045 | *None* | 12 | 132487182 | A | G | 0.13 | 1.09 | 0.08 | 0.20 | 0.11 | 0.45 | 0.06 |
| Nalls 2019 | rs9568188 | *CAB39L* | 13 | 49353596 | T | C | 0.35 | 1.01 | 0.01 | 0.75 | 0.63 | 0.26 | 0.06 |
| Nalls 2019 | rs4771268 | *None* | 13 | 97212767 | T | C | 0.40 | 1.05 | 0.05 | 0.25 | 0.65 | 0.79 | 0.07 |
| Nalls 2019 | rs12147950 | *MIPOL1* | 14 | 37520065 | T | C | 0.46 | 0.99 | -0.01 | 0.75 | 0.51 | 0.56 | -0.05 |
| Nalls 2019 | rs11158026 | *GCH1* | 14 | 54882151 | C | T | 0.46 | 1.23 | 0.21 | 0.00 | 0.55 | 0.31 | -0.08 |
| Nalls 2019 | rs3742785 | *RPS6KL1* | 14 | 74906331 | C | A | 0.20 | 0.98 | -0.02 | 0.76 | 0.20 | 0.23 | 0.07 |
| Nalls 2019 | rs979812 | *None* | 14 | 87997920 | G | T | 0.35 | 0.98 | -0.02 | 0.67 | 0.61 | 0.42 | 0.06 |
| Nalls 2019 | rs2251086 | *AC018618.1* | 15 | 61705186 | T | C | 0.15 | 0.88 | -0.13 | 0.03 | 0.81 | 0.84 | -0.12 |
| Nalls 2019 | rs6497339 | *SYT17* | 16 | 19266171 | A | T | 0.01 | 1.27 | 0.24 | 0.19 | 0.99 | 0.54 | 0.06 |
| Nalls 2019 | rs2904880 | *CD19/RABEP2* | 16 | 28933075 | C | G | 0.08 | 1.06 | 0.06 | 0.46 | 0.90 | 0.73 | -0.07 |
| Nalls 2019 | rs11150601 | *SETD1A* | 16 | 30966478 | G | A | 0.08 | 0.82 | -0.20 | 0.01 | 0.90 | 0.64 | 0.09 |

| | | | | | | | | | | | | | |
|---|---|---|---|---|---|---|---|---|---|---|---|---|---|
| Nalls 2019 | rs6500328 | *NOD2* | 16 | 50702745 | G | A | 0.20 | 1.00 | 0.00 | 0.96 | 0.18 | 0.43 | 0.06 |
| Nalls 2019 | rs3104783 | *CASC16* | 16 | 52602330 | C | A | 0.21 | 0.91 | -0.09 | 0.08 | 0.80 | 0.43 | 0.07 |
| Nalls 2019 | rs10221156 | *PHBP21* | 16 | 52935514 | A | G | 0.04 | 1.09 | 0.08 | 0.47 | 0.03 | 0.08 | -0.12 |
| Nalls 2019 | rs12600861 | *CHRNB1* | 17 | 7452302 | C | A | 0.23 | 1.04 | 0.04 | 0.42 | 0.22 | 0.34 | -0.06 |
| Nalls 2019 | rs12951632 | *RETREG3* | 17 | 42588995 | C | T | 0.44 | 0.94 | -0.06 | 0.14 | 0.45 | 0.27 | 0.06 |
| Nalls 2019 | rs2269906 | *UBTF* | 17 | 44216969 | A | C | 0.23 | 1.06 | 0.06 | 0.25 | 0.75 | 0.32 | 0.06 |
| Nalls 2019 | rs850738 | *FAM171A2* | 17 | 44357262 | G | A | 0.48 | 1.05 | 0.05 | 0.29 | 0.52 | 0.41 | -0.07 |
| Nalls 2019 | rs62053943 | *LINC02210-CRHR1* | 17 | 45666837 | NA | NA | NA | NA | NA | NA | 0.00 | 0.17 | -0.27 |
| Nalls 2019 | rs117615688 | *LINC02210-CRHR1* | 17 | 45720942 | NA | NA | NA | NA | NA | NA | None | 0.07 | -0.23 |
| Nalls 2019 | rs11658976 | *WNT3* | 17 | 46789439 | A | G | 0.38 | 0.99 | -0.01 | 0.81 | 0.40 | 0.62 | -0.06 |
| Nalls 2019 | rs61169879 | *BRIP1* | 17 | 61840005 | C | T | 0.44 | 0.98 | -0.02 | 0.62 | 0.56 | 0.15 | 0.08 |
| Nalls 2019 | rs666463 | *DNAH17* | 17 | 78429399 | T | A | 0.04 | 0.94 | -0.06 | 0.60 | 0.02 | 0.18 | 0.08 |
| Nalls 2019 | rs1941685 | *ASXL3* | 18 | 33724354 | G | T | 0.10 | 0.95 | -0.05 | 0.45 | 0.88 | 0.51 | 0.05 |
| Nalls 2019 | rs12456492 | *RIT2* | 18 | 43093415 | G | A | 0.40 | 1.13 | 0.12 | 0.01 | 0.36 | 0.34 | -0.10 |
| Nalls 2019 | rs8087969 | *None* | 18 | 51157219 | G | T | 0.28 | 0.94 | -0.07 | 0.17 | 0.24 | 0.44 | -0.06 |
| Nalls 2019 | rs55818311 | *SPPL2B/AC005258.1* | 19 | 2341049 | T | C | 0.30 | 0.98 | -0.02 | 0.68 | 0.34 | 0.67 | -0.07 |
| Nalls 2019 | rs77351827 | *CRLS1* | 20 | 6025395 | NA | NA | NA | NA | NA | NA | 0.00 | 0.13 | 0.08 |

| | | | | | | | | | | | | | |
|---|---|---|---|---|---|---|---|---|---|---|---|---|---|
| Nalls 2019 | rs2248244 | *DYRK1A* | 21 | 37480059 | A | G | 0.38 | 1.11 | 0.11 | 0.02 | 0.37 | 0.26 | 0.07 |
| Foo 2020 | rs6679073 | *PARK16* | 1 | 205787356 | A | C | 0.41 | 1.13 | 0.12 | 4.9E-03 | 0.50 | 0.25 | 0.21 |
| Foo 2020 | rs16846351 | *ITPKB* | 1 | 226659011 | G | C | 0.07 | 1.29 | 0.25 | 2.5E-03 | 0.07 | None | 0.29 |
| Foo 2020 | rs12278023 | *DLG2* | 11 | 83799074 | C | T | 0.49 | 0.93 | -0.07 | 7.8E-02 | 0.54 | 0.45 | -0.13 |
| Foo 2020 | rs141336855 | *LRRK2* | 12 | 39993947 | T | G | 0.03 | 1.58 | 0.46 | 1.9E-04 | 0.02 | None | 0.69 |
| Foo 2020 | rs4130047 | *RIT2* | 18 | 43098270 | C | T | 0.39 | 1.13 | 0.12 | 5.9E-03 | 0.36 | 0.34 | 0.13 |
| Foo 2020 | rs2292056 | *MCCC1* | 3 | 183017423 | G | T | 0.43 | 0.76 | -0.27 | 8.5E-10 | 0.57 | 0.20 | -0.19 |
| Foo 2020 | rs3816248 | *SCARB2/FAM47E* | 4 | 76179915 | C | T | 0.36 | 0.97 | -0.04 | 4.3E-01 | 0.33 | 0.13 | -0.14 |
| Foo 2020 | rs6826785 | *SNCA* | 4 | 89761323 | C | T | 0.43 | 1.32 | 0.28 | 1.6E-10 | 0.54 | 0.07 | 0.29 |
| Foo 2020 | rs246814 | *SV2C* | 5 | 76303383 | T | C | 0.09 | 1.11 | 0.11 | 1.5E-01 | 0.09 | 0.08 | 0.22 |
| Foo 2020 | rs1887316 | *FYN* | 6 | 111830249 | A | G | 0.12 | 0.82 | -0.20 | 1.8E-03 | 0.12 | 0.11 | -0.19 |
| Foo 2020 | rs9638616 | *WBSCR17/GALNT17* | 7 | 71285507 | T | G | 0.46 | 0.99 | -0.01 | 8.7E-01 | 0.48 | 0.32 | 0.13 |

### Supplementary Table 4. Polygenic Risk Score (PRS) Variants and Weights

This table summarizes the variants included in the construction of the polygenic risk scores (PRS) evaluated in this study.

**Supplementary Table 4A.**
Seventy-six single-nucleotide polymorphisms (SNPs) derived from the **European-based PRS** were included as the baseline model, based on the summary statistics reported by *Nalls et al., 2019*. The corresponding effect alleles and weights are listed in Supplementary Table 3A.

**Supplementary Table 4B.**
Eleven **Asian-specific SNPs** were incorporated according to *Foo et al., 2020*. Nine overlapping loci between the European and Asian datasets were removed from the baseline list before model integration.

**Supplementary Table 4C.**
Four additional SNPs were included to reflect **Taiwan-specific genetic architecture**, comprising two **LRRK2 exonic variants** (p.N551K and p.R1398H) and two novel variants located in **VPS13C and SCARB2 identified in this study.**

**Supplementary Table 4A.**

| rsID | chromosome | position (hg38) | effect allele | effect weight |
|---|---|---|---|---|
| rs6658353 | 1 | 161499264 | C | 0.065 |
| rs11578699 | 1 | 171750629 | T | -0.0704 |
| rs823118 | 1 | 205754444 | T | 0.1066 |
| rs11557080 | 1 | 205768611 | A | 0.1315 |
| rs4653767 | 1 | 226728377 | T | 0.0833 |
| rs10797576 | 1 | 232528865 | T | 0.1114 |
| rs2042477 | 2 | 95335195 | A | -0.0657 |
| rs11683001 | 2 | 101780501 | A | 0.0705 |
| rs57891859 | 2 | 134707046 | A | 0.0807 |
| rs1474055 | 2 | 168253884 | T | 0.1796 |
| rs6808178 | 3 | 28664199 | T | 0.0658 |
| rs12497850 | 3 | 48711556 | T | 0.0636 |
| rs55961674 | 3 | 122478045 | T | 0.0861 |
| rs11707416 | 3 | 151391177 | A | -0.0627 |
| rs1450522 | 3 | 161359842 | A | -0.0616 |
| rs10513789 | 3 | 183042285 | T | 0.1485 |

| | | | | |
|---|---|---|---|---|
| rs34311866 | 4 | 958159 | T | -0.2126 |
| rs4698412 | 4 | 15735725 | A | 0.1035 |
| rs34025766 | 4 | 17967188 | A | -0.0839 |
| rs6825004 | 4 | 76189212 | C | 0.0622 |
| rs4101061 | 4 | 76226816 | A | -0.0912 |
| rs6854006 | 4 | 76276901 | T | -0.0912 |
| rs356182 | 4 | 89704960 | A | -0.2774 |
| rs13117519 | 4 | 113447909 | T | 0.0875 |
| rs62333164 | 4 | 169662006 | A | -0.0638 |
| rs1867598 | 5 | 60842132 | A | -0.1554 |
| rs26431 | 5 | 103030090 | C | 0.0621 |
| rs11950533 | 5 | 134863415 | A | -0.0916 |
| rs4140646 | 6 | 27771022 | A | 0.0833 |
| rs9261484 | 6 | 30140906 | T | -0.0635 |
| rs112485576 | 6 | 32610995 | A | -0.1676 |
| rs12528068 | 6 | 71778059 | T | 0.0657 |
| rs997368 | 6 | 111922088 | A | 0.0714 |
| rs75859381 | 6 | 132889222 | T | -0.2207 |
| rs199351 | 7 | 23260430 | A | 0.1016 |
| rs76949143 | 7 | 66544864 | A | -0.1432 |
| rs620513 | 8 | 16840084 | T | -0.0856 |
| rs2280104 | 8 | 22668467 | T | 0.0556 |
| rs2086641 | 8 | 129889663 | T | -0.0605 |
| rs13294100 | 9 | 17579692 | T | -0.0859 |
| rs10756907 | 9 | 17727067 | A | -0.0926 |
| rs6476434 | 9 | 34046393 | T | -0.0615 |
| rs896435 | 10 | 15515407 | T | 0.0735 |
| rs10748818 | 10 | 102255522 | A | -0.079 |
| rs7938782 | 11 | 10537230 | A | 0.087 |
| rs12283611 | 11 | 83776234 | A | -0.0645 |

| | | | | |
|---|---|---|---|---|
| rs3802920 | 11 | 133917106 | T | 0.1073 |
| rs76904798 | 12 | 40220632 | T | 0.1439 |
| rs7134559 | 12 | 46025303 | T | -0.0539 |
| rs10847864 | 12 | 122842051 | T | 0.1478 |
| rs11610045 | 12 | 132487182 | A | 0.0601 |
| rs9568188 | 13 | 49353596 | T | 0.0617 |
| rs4771268 | 13 | 97212767 | T | 0.0675 |
| rs12147950 | 14 | 37520065 | T | -0.0529 |
| rs11158026 | 14 | 54882151 | T | -0.0842 |
| rs3742785 | 14 | 74906331 | A | 0.0707 |
| rs979812 | 14 | 87997920 | T | 0.061 |
| rs2251086 | 15 | 61705186 | T | -0.1186 |
| rs6497339 | 16 | 19266171 | A | 0.063 |
| rs2904880 | 16 | 28933075 | C | -0.065 |
| rs11150601 | 16 | 30966478 | A | 0.0907 |
| rs6500328 | 16 | 50702745 | A | 0.0586 |
| rs3104783 | 16 | 52602330 | A | 0.0668 |
| rs10221156 | 16 | 52935514 | A | -0.1156 |
| rs12600861 | 17 | 7452302 | A | -0.0565 |
| rs12951632 | 17 | 42588995 | T | 0.0642 |
| rs2269906 | 17 | 44216969 | A | 0.0631 |
| rs850738 | 17 | 44357262 | A | -0.071 |
| rs11658976 | 17 | 46789439 | A | -0.0624 |
| rs61169879 | 17 | 61840005 | T | 0.082 |
| rs666463 | 17 | 78429399 | A | 0.076 |
| rs1941685 | 18 | 33724354 | T | 0.0531 |
| rs12456492 | 18 | 43093415 | A | -0.0983 |
| rs8087969 | 18 | 51157219 | T | -0.0578 |
| rs55818311 | 19 | 2341049 | T | -0.0696 |
| rs2248244 | 21 | 37480059 | A | 0.0714 |

**Supplementary Table 4B**

| rsID | chromosome | position (hg38) | effect allele | effect weight |
|---|---|---|---|---|
| rs6679073 | 1 | 205787356 | A | 0.21 |
| rs16846351 | 1 | 226659011 | G | 0.29 |
| rs12278023 | 11 | 83799074 | C | -0.13 |
| rs141336855 | 12 | 39993947 | T | 0.69 |
| rs4130047 | 18 | 43098270 | C | 0.13 |
| rs2292056 | 3 | 183017423 | G | -0.19 |
| rs3816248 | 4 | 76179915 | C | -0.14 |
| rs6826785 | 4 | 89761323 | C | 0.29 |
| rs246814 | 5 | 76303383 | T | 0.22 |
| rs1887316 | 6 | 111830249 | A | -0.19 |
| rs9638616 | 7 | 71285507 | T | 0.13 |

**Supplementary Table 4C**

| rsID | chromosome | position | effect_allele | effect_weight |
|---|---|---|---|---|
| rs34778348 | 12 | 40363526 | A | 0.47 |
| rs33949390 | 12 | 40320043 | C | 0.398 |
| rs76591264 | 4 | 76217199 | G | 0.183 |
| rs12900645 | 15 | 61994009 | G | -0.189 |

**Supplementary Table 5. Polygenic Risk Score (PRS) Stratification**

This table presents the odds ratios (ORs) and 95% confidence intervals (CIs) for Parkinson's disease (PD) risk across ten deciles of the polygenic risk score (PRS) distribution in the Taiwanese cohort.
The first decile (D1) served as the reference group, representing individuals with the lowest PRS values. ORs were calculated relative to D1, adjusting for age, sex, and the top ten principal components of ancestry. A progressive increase in PD risk was observed across higher PRS deciles, with the tenth decile (D10) showing the highest risk enrichment.

| Decile | OR | CI_low | CI_high | estimate | std.error | statistic | p.value | conf.low | conf.high |
|---|---|---|---|---|---|---|---|---|---|
| D1 | 1 | 1 | 1 | NA | NA | NA | NA | NA | NA |
| D2 | 1.52 | 1.16 | 2.00 | 1.52 | 0.14 | 3.02 | 2.5E-03 | 1.16 | 2.00 |
| D3 | 1.34 | 1.02 | 1.76 | 1.34 | 0.14 | 2.08 | 3.7E-02 | 1.02 | 1.76 |
| D4 | 1.80 | 1.38 | 2.36 | 1.80 | 0.14 | 4.27 | 1.9E-05 | 1.38 | 2.36 |
| D5 | 2.01 | 1.53 | 2.63 | 2.01 | 0.14 | 5.05 | 4.4E-07 | 1.53 | 2.63 |
| D6 | 2.19 | 1.67 | 2.87 | 2.19 | 0.14 | 5.69 | 1.2E-08 | 1.67 | 2.87 |
| D7 | 2.29 | 1.75 | 3.00 | 2.29 | 0.14 | 6.01 | 1.8E-09 | 1.75 | 3.00 |
| D8 | 2.74 | 2.09 | 3.60 | 2.74 | 0.14 | 7.28 | 3.3E-13 | 2.09 | 3.60 |
| D9 | 3.23 | 2.46 | 4.25 | 3.23 | 0.14 | 8.41 | 4.0E-17 | 2.46 | 4.25 |
| D10 | 4.70 | 3.56 | 6.25 | 4.70 | 0.14 | 10.81 | 3.1E-27 | 3.56 | 6.25 |

**Supplementary Figure S1.** Principal component analysis of the 1000 Genomes reference panel with projection of the Taiwanese cohort (TW, gray). TW samples cluster with the East Asian (EAS, green) superpopulation on PC1–PC2, consistent with East Asian ancestry. PCs were computed on LD-pruned autosomal SNPs in the 1000 Genomes Project; Taiwanese samples were projected without refitting. Other superpopulations are shown for context (AFR, AMR, EUR, SAS).

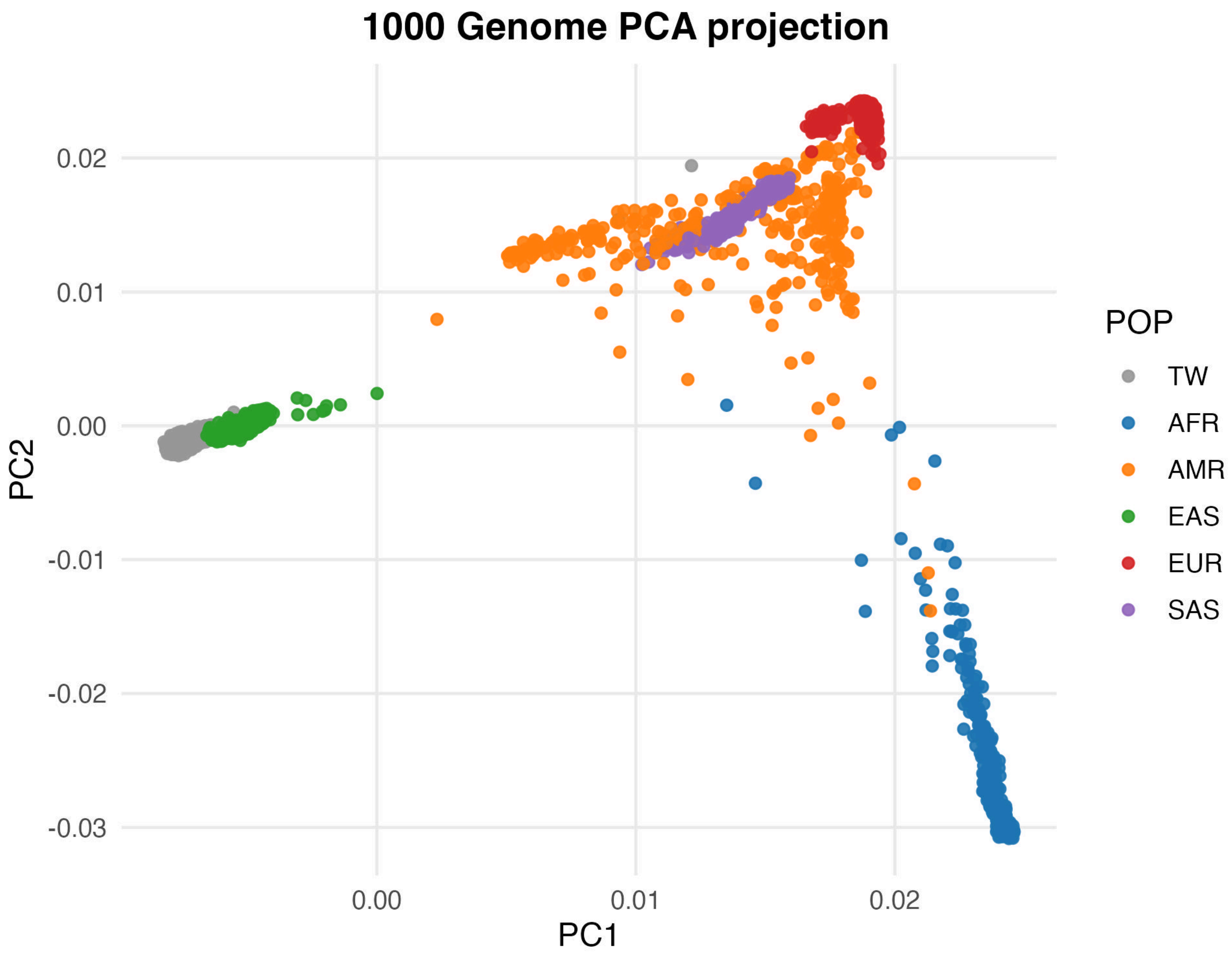

**Supplementary Figure S2. Quantile–quantile (QQ) plot of genome-wide association p-values.**

The QQ plot demonstrates excellent calibration of test statistics, with a genomic control inflation factor ($\lambda_{GC}$ = 1.02). The observed p-value distribution closely follows the expected null distribution (dashed line), indicating minimal residual population stratification or technical artifacts.

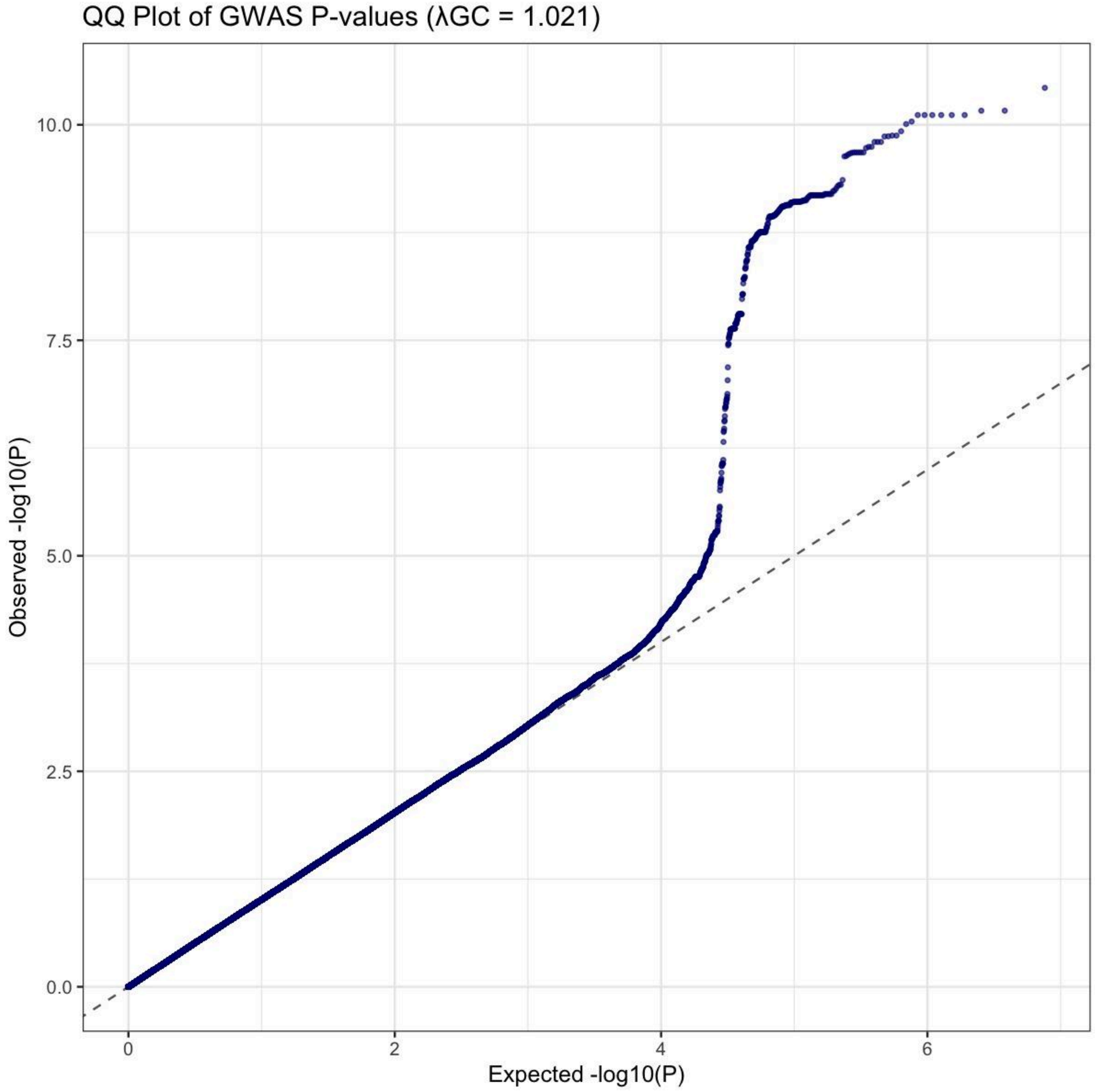

**Supplementary Figure S3. Regional association plots for key loci (a–h).** LocusZoom-style plots show $-\log_{10}(P)$ by genomic position (hg38) across ±300 kb around each lead SNP. LD is computed from cohort controls; lead variants are highlighted in purple and other variants are colored by $r^2$ with the lead.

(a) **MCCC1** (lead rs34822404, chr3:183,028,420); (b) **GCH1** (lead rs3783640, chr14:54,885,803); (c) **PPARGC1A** (lead rs2970847, chr4:23,756,312); (d) **GALNT13** (lead rs10186542, chr2:154,088,802); (e) **VPS13C** (lead rs12900645, chr15:61,994,009), distant from and not in LD with rs2251086 reported by Nalls; rs2251086 lies within *VPS13C*, supporting *VPS13C* as the risk locus; (f) **SCARB2** (lead rs76591264, chr4:76,217,199), not in LD with rs4101061 reported by Nalls.

**a. MCCC1** (lead rs34822404, chr3:183,028,420).

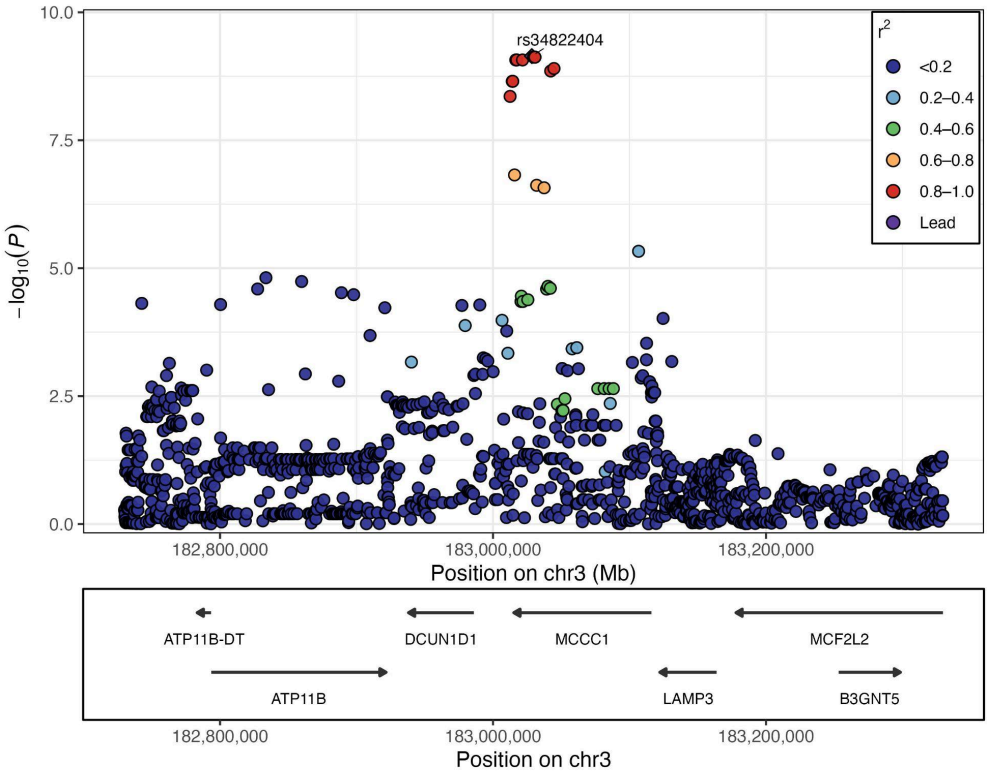

b**. GCH1** (lead rs3783640, chr14:54,885,803).

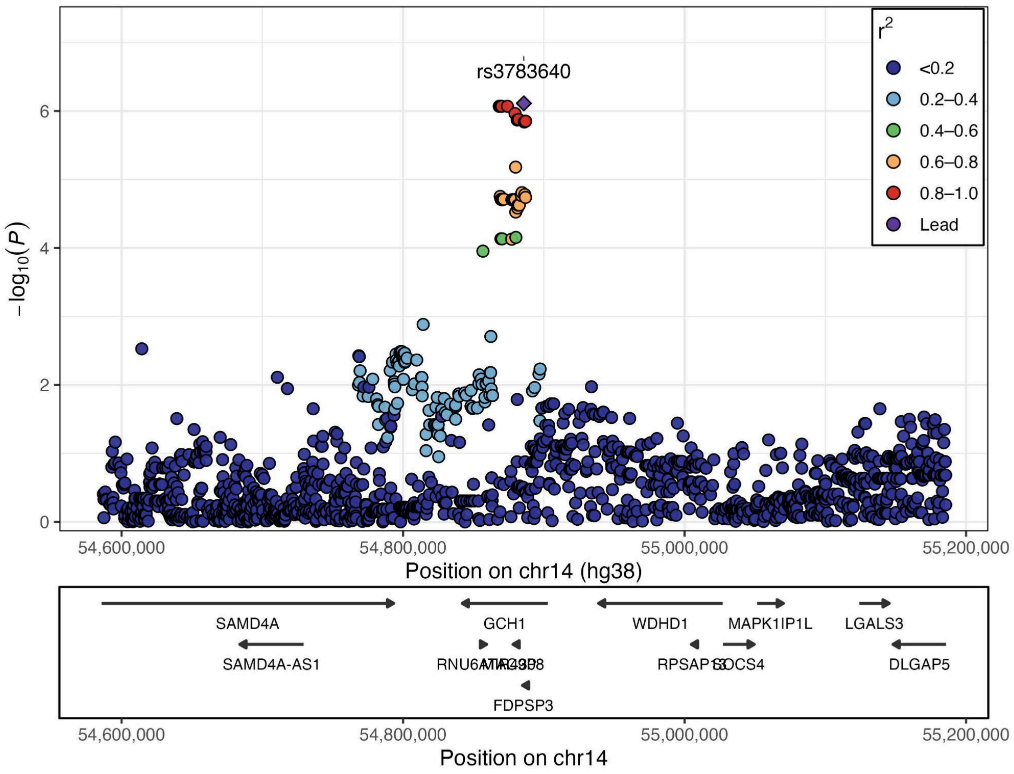


**c. PPARGC1A** (lead rs2970847, chr4:23,756,312).

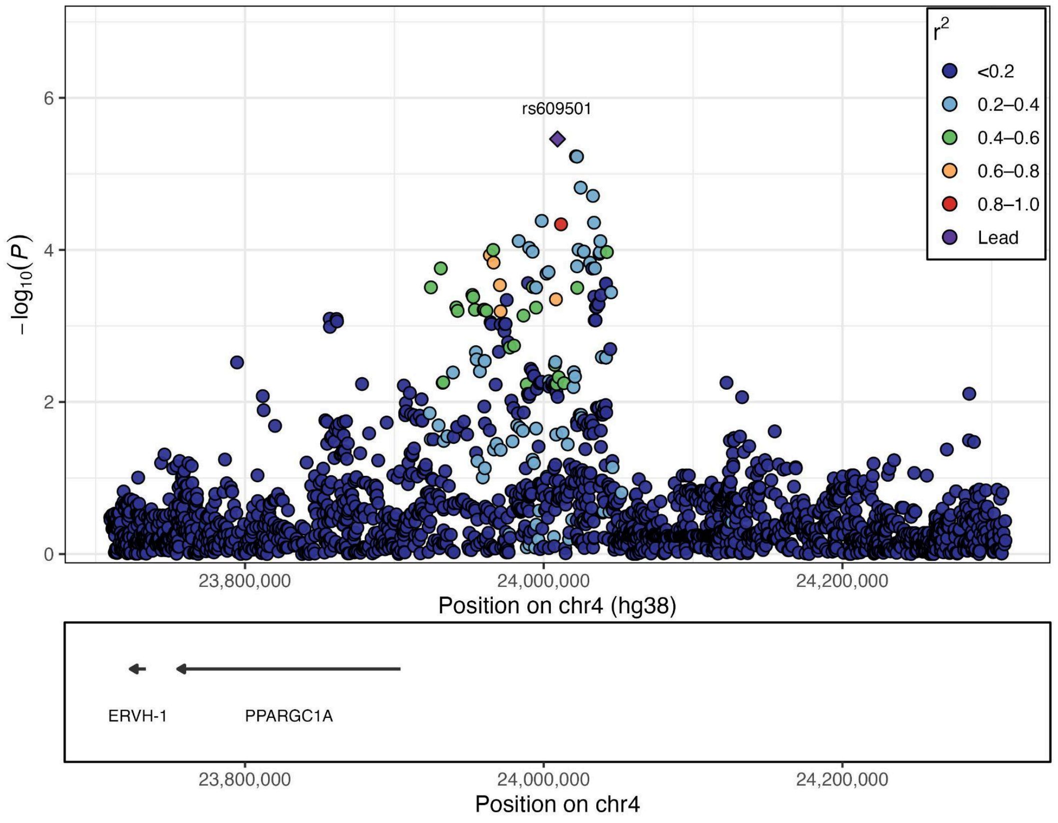

**d. GALNT13** (lead rs10186542, chr2:154,088,802).

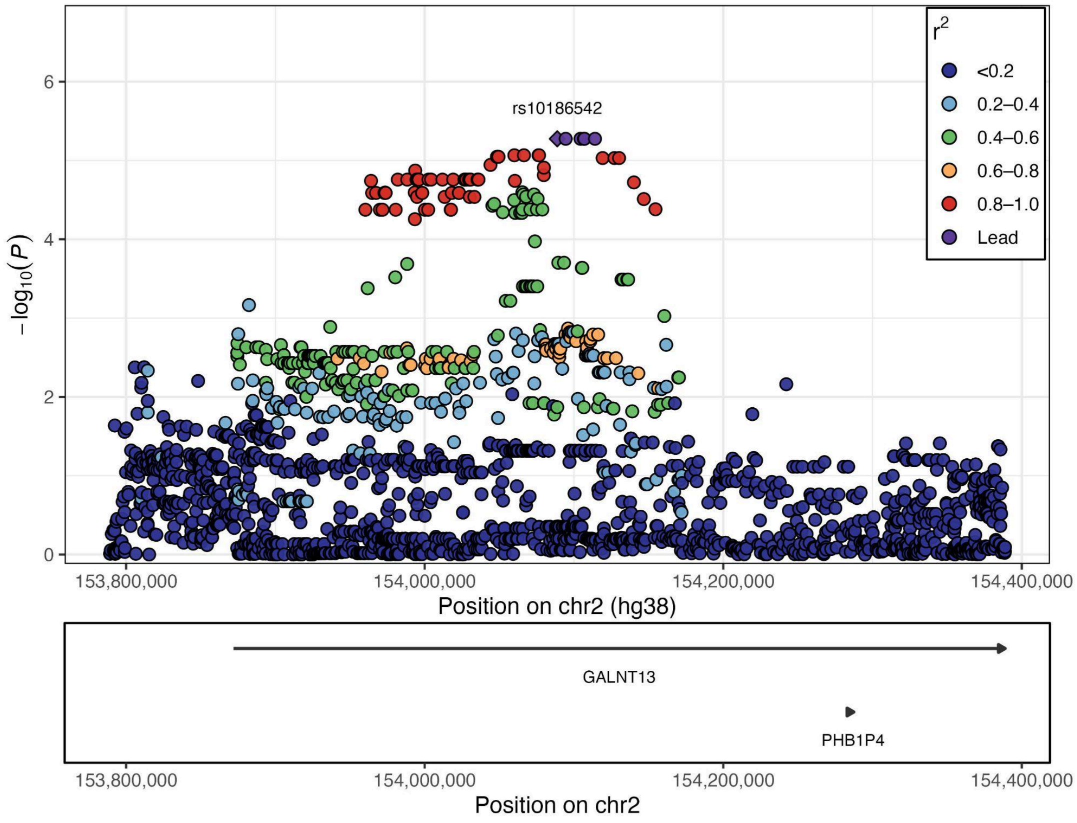


**e. VPS13C** (lead rs12900645, chr15:61,994,009). This signal is distant from and not in LD with rs2251086 reported by Nalls et al.; rs12900645 lies within *VPS13C*, supporting *VPS13C* as the risk locus.

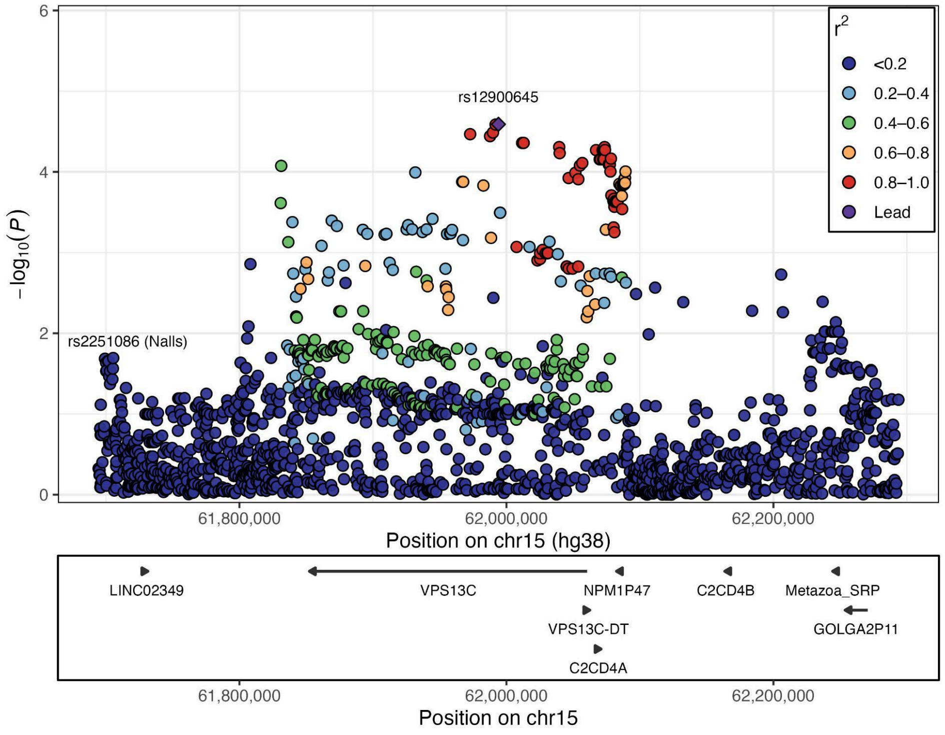

**f. SCARB2** (lead rs76591264, chr4:76,217,199). This signal is not in LD with rs4101061 reported by Nalls et al.

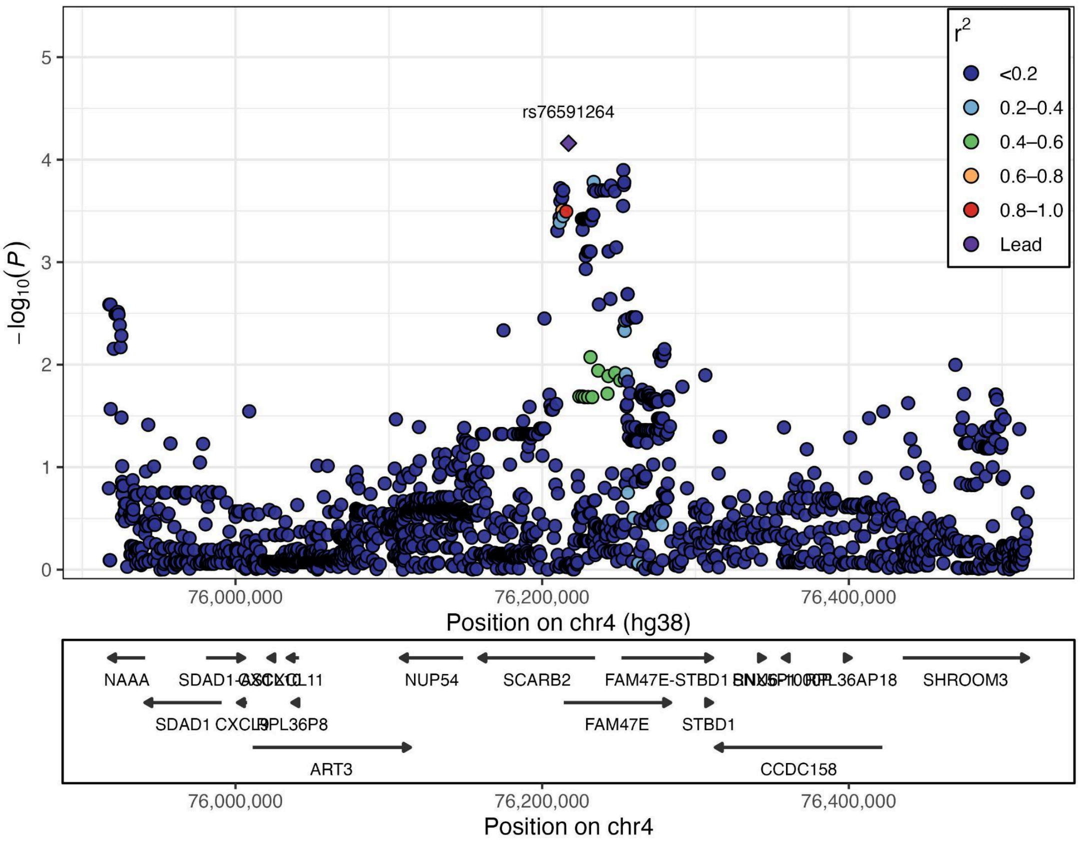

**Supplementary Figure S4. Haplotype structure and association of *GCH1* in the Taiwanese Parkinson's disease cohort.**

(a) Lower-triangle LD heatmap among rs841, rs11158026, and rs3783640 (hg38), calculated in cohort controls; the Taiwan-specific rs3783640 is in LD with rs11158026 (Nalls et al.), and both are in partial LD with rs841 ($r^2 = 0.31$). (b) Haplotype composition and frequencies (displayed for haplotypes with control frequency >1%), ordered by control frequency. (c) Forest plot of haplotype associations with PD (OR, 95% CI). Haplotypes carrying rs3783640-C and rs11158026-T show a protective effect against PD, whereas rs841 alone is not significantly associated.

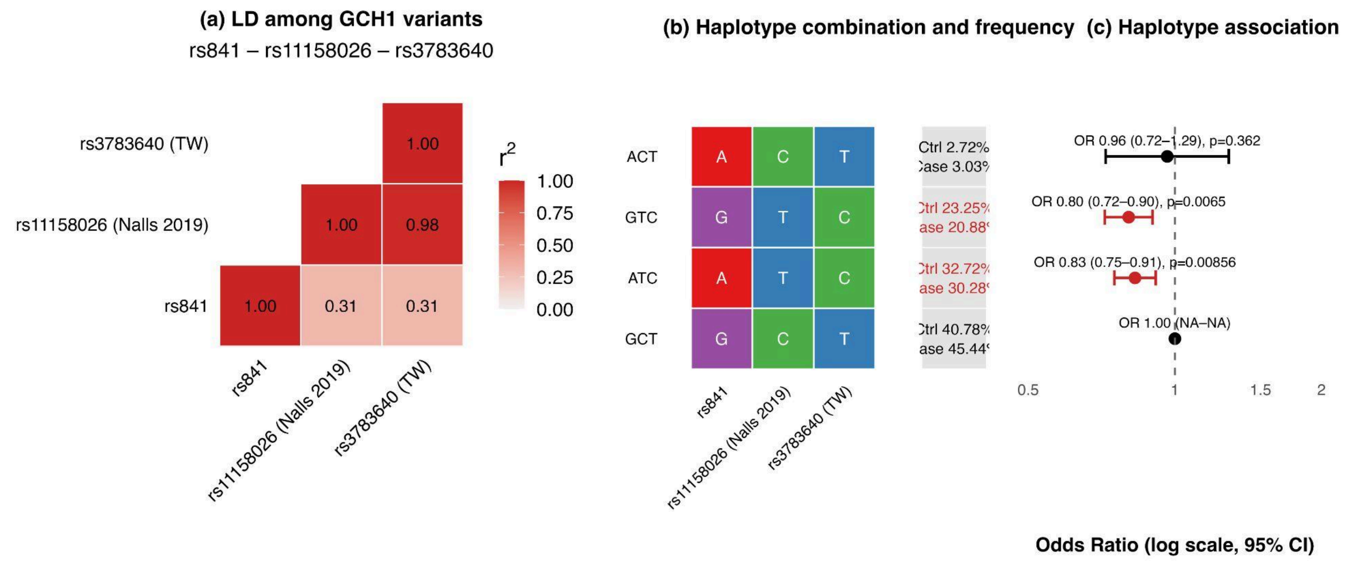
